# A microbiome-directed therapeutic food for children recovering from severe acute malnutrition

**DOI:** 10.1101/2024.06.11.24307076

**Authors:** Steven J. Hartman, Matthew C. Hibberd, Ishita Mostafa, Naila N. Nahar, Md. Munirul Islam, Mahabub Uz Zaman, Sayeeda Huq, Mustafa Mahfuz, Md. Tazul Islam, Kallol Mukherji, Vaha Akbary Moghaddam, Robert Y. Chen, Michael A. Province, Daniel M. Webber, Suzanne Henrissat, Bernard Henrissat, Nicolas Terrapon, Dmitry A. Rodionov, Andrei L. Osterman, Michael J. Barratt, Tahmeed Ahmed, Jeffrey I. Gordon

## Abstract

Severe acute malnutrition (SAM), defined anthropometrically as a weight-for-length z-score more than 3 standard deviations below the mean (WLZ<-3), affects 19 million children under 5-years-old worldwide. Complete anthropometric recovery after standard inventions is rare with children often left with moderate acute malnutrition (MAM; WLZ -2 to -3). Here we conduct a randomized controlled trial (RCT), involving 12-18-month-old Bangladeshi children from urban and rural sites, who after hospital-based treatment for SAM received a 3-month intervention with a microbiota-directed complementary food (MDCF-2) or a ready-to-use supplementary food (RUSF) as they transitioned to MAM. The rate of WLZ improvement was significantly greater with MDCF-2 than the more calorically-dense RUSF, as we observed in a previous RCT of Bangladeshi children with MAM without antecedent SAM. A correlated meta-analysis of aptamer-based measurements of 4,520 plasma proteins in this and the prior RCT revealed 215 proteins positively-associated with WLZ (prominently those involved in musculoskeletal and CNS development) and 44 negatively-associated proteins (related to immune activation), with a significant enrichment in levels of the positively WLZ-associated proteins in the MDCF-2 arm. Characterizing changes in 754 bacterial metagenome-assembled genomes in serially collected fecal samples disclosed the effects of acute rehabilitation for SAM on the microbiome, its transition as each child achieves a state of MAM, and how specific strains of *Prevotella copri* function at the intersection between MDCF-2 glycan metabolism and the rescue of growth faltering. These results provide a rationale for further testing the generalizability of the efficacy of MDCF and identify biomarkers for defining treatment responses.

**One Sentence Summary:** A microbiota-directed food that targets specific gut bacterial taxa promotes growth in Bangladeshi children recovering from severe acute malnutrition

## INTRODUCTION

Childhood undernutrition is a global health challenge that is not due to food insecurity alone. Its sequelae include long-term defects in linear growth (stunting), disturbed metabolism, and immune function, as well as perturbed neurodevelopment - chronic conditions that have proven to be largely resistant to current therapeutic interventions and pose significant societal burdens in low- and middle-income countries (*1*).

Severe acute malnutrition (SAM) in non-edematous children is defined by either a WLZ score more than three standard deviations below the median value for a multi-national World Health Organization (WHO) cohort of comparably-aged healthy infants and children or a mid-upper arm circumference of <115 mm. SAM accounts for 23% of all cases of acute malnutrition in children under five, with the remaining 77% of children classified as having moderate acute malnutrition (MAM; WLZ - 2 to -3)(*1*). Analyses of the fecal microbiota of children with MAM or SAM have revealed that community assembly is perturbed, resulting in microbiota configurations that resemble those of chronologically younger children (*2*). This microbiota immaturity is not fully repaired with commonly used therapeutic interventions, leaving these children with a persistent defect in the developmental biology of their microbial communities (*2*–*4*). In preclinical tests designed to distinguish cause from effect, gnotobiotic mice were colonized with fecal microbiota samples collected from healthy children or from chronologically age-matched children with acute malnutrition; microbial communities from the latter transmitted impaired weight gain and altered bone growth phenotypes, plus immune and metabolic abnormalities (*3, 5, 6*). A machine learning-based analysis of these transplanted communities revealed bacterial strains whose representation correlated with various facets of growth, including lean body-mass gain (*5*).

Efforts to develop affordable, culturally acceptable interventions for repairing the microbiota of undernourished children began by screening combinations of Bangladeshi complementary foods (foods given as children are being weaned) in gnotobiotic mice and gnotobiotic piglets harboring gut communities from undernourished Bangladeshi children. These efforts yielded microbiota-directed complementary food (MDCF) prototypes that altered the abundances and expressed metabolic activities of bacterial strains that are inappropriately represented in immature microbiota (*3*). In a 3-month randomized controlled clinical trial (RCT) involving 12-18-month-old Bangladeshi children with MAM living in an urban slum (Mirpur) in Dhaka, supplementation with a lead MDCF prototype (MDCF-2) produced a significantly greater rate of ponderal growth (β-WLZ) compared to a standard ready-to-use supplementary food (RUSF), even though MDCF-2 had 15% fewer calories per serving (*7*). MDCF-2 contained four complementary food components (chickpea flour, soybean flour, peanut paste, green banana) plus soybean oil, sugar and micronutrients (*3, 7*). The superior effect of MDCF-2 on β-WLZ was accompanied by more complete microbiota repair plus greater increases in levels of plasma protein mediators and biomarkers of musculoskeletal and central nervous system (CNS) development at the end of treatment (*7*) and a significant improvement in linear growth trajectories in the 2-year period after cessation of treatment (*8*). Shotgun sequencing of DNA isolated from fecal biospecimens serially collected during this trial identified 75 bacterial metagenome assembled genomes (MAGs) whose abundances were significantly positively associated with WLZ and whose genomes were enriched for metabolic pathways involved in utilization of plant polysaccharides (*9*). Combining microbial RNA-Seq with mass spectrometry of MDCF-2 carbohydrates highlighted two *Prevotella copri* MAGs (strains), positively associated with WLZ, that were principal contributors to MDCF-2-induced expression of metabolic pathways involved in utilization of its component glycans (*9*). The genomes of several *P. copri* strains that were subsequently cultured from the Bangladeshi study population closely resembled those of the two MAGs, including similarities in their polysaccharide utilization loci (PULs). The predicted specificities of carbohydrate active enzymes (CAZymes) expressed by PULs in the *P. copri* MAGs correlated with (i) the *in vitro* growth rates of these *P. copri* strains in the presence of purified glycans representing structures present in MDCF-2 and (ii) the levels of breakdown products of MDCF-2 glycans identified in the feces of trial participants. Follow-up *‘*reverse translation’ experiments, involving a gnotobiotic mouse model of postnatal microbiota development colonized with collections of age-discriminatory and WLZ-associated bacterial strains cultured from the Bangladeshi study population have provided direct evidence that *P. copri* plays a key role in mediating MDCF-2 metabolism, intestinal epithelial cell metabolism and weight gain (*10*).

Hospital-based protocols for acute rehabilitation of children >6 months of age with SAM typically involve a short course of parenteral and oral antibiotics, and nutritional resuscitation with fortified milk formulations and/or complementary foods. A recent meta-analysis suggests that after initial treatment, these children often transition to a state of persistent MAM (‘post-SAM MAM’), with anthropometric status after treatment being the strongest predictor of relapse (*11*). In the current study, we performed a randomized controlled study of the effects of MDCF-2 versus RUSF in 12-18-month-old Bangladeshi children who presented with non-edematous SAM and required hospitalization for their initial acute nutritional resuscitation. Participants lived either in the Mirpur urban slum or in Kurigram, a rural district in northern Bangladesh. The objectives of the study were to (i) determine whether the superior rate of change in WLZ obtained with MDCF-2 supplementation of children with ‘primary’ MAM also occurs in children with ‘post-SAM’ MAM, (ii) determine how the gut microbiome and plasma proteome respond to standard protocols for acute nutritional resuscitation of SAM, and as the child subsequently transitions to a state of post-SAM MAM, and (iii) ascertain the extent to which WLZ-associated MAGs and host proteomic responses to treatment are shared between similarly-aged children with primary MAM and post-SAM MAM and differentially affected by MDCF-2 compared to RUSF. As such, this study can be viewed as a test of the generalizability of the efficacy and mechanism of action of MDCF-2 in acutely malnourished children.

## RESULTS

### Study design

The primary outcome measure for this randomized, parallel assignment clinical trial was the effect of MDCF-2 versus RUSF treatment on the rate of ponderal growth (β-WLZ), while the secondary outcomes were treatment-associated changes in the plasma proteome and the fecal microbiome in children with post-SAM MAM (referred to hereafter as ‘MAM’ unless specified otherwise). The study design is described in **Fig. 1A**. A total of 124 children aged 12-18 months [14.7 ± 2.3 (mean ± SD)] with non-edematous SAM were enrolled (n=50/treatment group at the Kurigram site, n=12/treatment group at the Mirpur site) (see **table S1** for enrollment characteristics). At enrollment, trial participants received a standard management protocol for stabilization/acute rehabilitation developed for treating hospitalized children with ‘complicated’ SAM (i.e., those with concomitant severe infections and/or dehydration). This regimen included an intravenous antibiotic (gentamicin) as well as oral antibiotics (ampicillin, amoxicillin), plus nutritional rehabilitation (F-75 followed by F-100 therapeutic milks, with concomitant administration of rice- and lentil-based Khichuri Halwa) (**Fig. 1B**) (*12*). After attaining a WLZ score above -3, signifying a transition from SAM to MAM, participants were randomly assigned to receive either MDCF-2 or RUSF for 3 months. The treatment protocol used was the same as in our previous RCT of children with primary MAM (*7*); 25g servings of either therapeutic food were administered twice a day (under supervision by icddr,b field staff), thus providing ∼220-250 kcal/day, an amount intended to satisfy ∼25% of each child’s daily caloric requirements. After completing the 3-month intervention, children were followed for an additional month (**Fig. 1A**). Breastfeeding was encouraged throughout all phases of the study as per WHO recommendations.

**Fig. 1.**
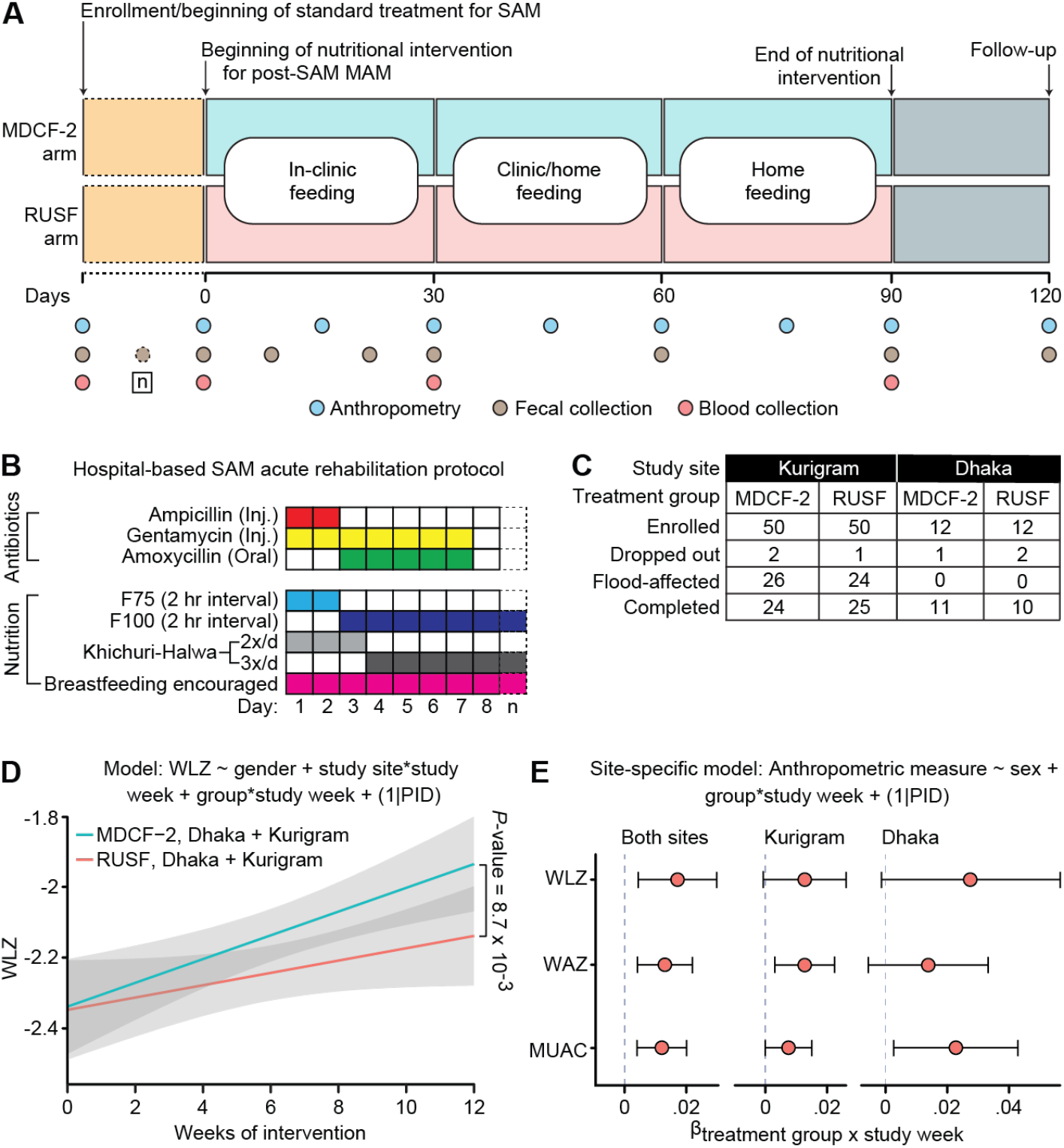
Study design and anthropometric analyses across sites and treatment groups. **(A)** Study design. (**B**) Hospital-based acute nutritional rehabilitation protocol for SAM. **(C)** Accounting of participants enrolled, dropped out, flood affected, and completing the study in Dhaka and Kurigram. Only children residing in Kurigram were affected by flooding during the trial. (**D**) Results of linear mixed effects modeling of the study-wide effect of MDCF-2 versus RUSF treatment on the primary outcome measure - rate of change of WLZ over time. Flood-affected participants were excluded from this analysis. The *P*-value corresponds to the effect of the term representing the ‘treatment group x study week’ interaction. (**E**) Results of modeling the primary outcome measure, plus additional anthropometric measures, across both sites (left), in Kurigram (middle), and in Dhaka (right). Each data point indicates the β-coefficient for the effect of the ‘treatment group x study week’ term on each of the indicated anthropometric measures. The upper and lower 95% confidence intervals for each coefficient estimate are indicated as error bars. The RUSF treatment group was used as the reference value; hence, positive coefficients indicate increases in anthropometric measurements associated with MDCF-2 treatment.

### Ponderal growth responses

A severe flood affected half of the trial participants residing in Kurigram, the primary study site, during the period of MDCF-2 or RUSF treatment (**fig. S1**). Although there was not a statistically significant difference between the baseline (pre-intervention) anthropometry of trial participants who would and would not be affected by this flood (*P* > 0.05, t-tests), the flood did have a significant impact on the primary outcome measure [*β*_WLZ_ = 0.017 (0.009, 0.014), *P* = 1.5×10^−5^, linear mixed effects modeling] despite efforts of the study team to provide trial-associated nutritional supplements and other support during this period of profound disruption and household relocation (**table S2**). Based on these findings, samples from children affected by the flood (n = 26 in the MDCF-2 arm and n = 24 in the RUSF arm) were excluded from our downstream multi-omic analyses, along with those from three children in each experimental arm who did not complete the study (**Fig. 1C, fig. S2**).

Acute rehabilitation for SAM resulted in each child achieving a WLZ > -3 within 7-13 days for those residing in Kurigram and within 7-19 days for those living in Mirpur. There was not a statistically significant relationship between the severity of SAM at enrollment (as measured by WLZ) and the length of time required for participants to exit the acute rehabilitation phase (Spearman’s rho = -0.08; *P* > 0.05); this was the case when participants at both study sites were considered together and when those at each study site were evaluated independently. Within each site, the anthropometric characteristics of children at baseline, just prior to the start of the MDCF-2 or RUSF nutritional intervention, were not significantly different (*P* > 0.05, t-test; **table S3**), although there were differences in anthropometry between the study sites [baseline WLZ: -2.56 ± 0.34 (Mirpur) versus -2.19 ± 0.38 (Kurigram); *P* = 3.26×10^−4^, t-test]. Moreover, compared to our previous study of 12-18-month-old Bangladeshi children with primary MAM (7), all anthropometric parameters were significantly lower prior to treatment in this population with post-SAM MAM (**table S4**).

Measurement of the weights of uneaten supplement after each feeding session disclosed that children consumed on average > 96% of the provided supplement across each arm, site, or sex, and there was not a statistically significant difference between the quantities of MDCF-2 and RUSF consumed (**table S5A**). There were also no significant differences in the proportion of children who satisfied WHO requirements for minimum meal frequency or minimum acceptable diet (assessed using food frequency questionnaires) during this period of treatment. Moreover, there was not a statistically significant difference in the amount of breast milk consumed between members of the two treatment groups (q-value > 0.05, **table S5B-E**).

The primary outcome measure of the effect of the MDCF-2 compared to the RUSF intervention on ponderal growth (β-WLZ) was modeled while controlling for sex, study site and repeated sampling of each individual. An intent-to-treat analysis that included all flood-affected and unaffected children revealed no significant differences in β-WLZ between the treatment groups (**table S6A**,**B**). However, focusing our analyses on flood-unaffected children across both study sites revealed that those who received MDCF-2 exhibited a significantly faster rate of ponderal growth compared to those who received RUSF (*β*_WLZ_ = 0.017 [0.004, 0.030], *P* = 8.73×10^−3^) (see Eq. 7 in *Methods*; **Fig. 1D,E, table S6C**). During the 3-month intervention, weight-for-age Z score (WAZ) and mid-upper arm circumference (MUAC) also improved at a significantly greater rate in the MDCF-2 compared to the RUSF group (**Fig. 1E)**. Consistent with our findings in children with primary MAM (*7*), there was not a statistically significant difference in the rate of linear growth between the two groups during treatment or during the 1-month follow-up period (*P* = 0.440) (**table S6D-F**). The occurrence of co-morbidities (fever, cough, diarrhea, or runny nose) during the MAM phase was not significantly associated with treatment type or with changes in WLZ (see *Methods*, **table S7**).

### Effects of treatment on the plasma proteome

We performed an aptamer-based proteomic analysis of plasma collected at four timepoints: when participants presented with SAM; at the completion of acute nutritional rehabilitation; after 1 month and at the end of the 3-month period of MDCF/RUSF treatment for MAM. Datasets from the current study were filtered to the 6,982 aptamers targeting 6,138 proteins that met quality control thresholds (see *Methods*). We had also used a subset of these aptamers (n=4,777) to measure levels of 4,520 of these plasma proteins in participants in the previous primary MAM RCT (*7*).

Datasets for proteins that were quantified in both studies were analyzed using two complementary approaches. The first involved the use of linear models to define the relationship between the change in abundance of each protein during the MAM phase of the current study to the change in WLZ during the same period, while controlling for study site, treatment group and sex (see *Methods*). Our first analysis tested for significant differences between treatment arms of the change in each protein’s level during MAM-phase treatment and did not identify any significant differences after applying false-discovery rate corrections (q-value > 0.1; Eq. 12; **table S8A**). We subsequently searched for proteins associated with ponderal growth responses and identified 112 proteins with statistically significant associations with WLZ during the MAM phase, of which 93 were positively associated (q-value < 0.05; see Eq. 15 in *Methods***; table S8B**). Gene Set Enrichment Analysis (GSEA) disclosed that compared to RUSF, MDCF-2 treatment resulted in a net increase in levels of proteins with a positive WLZ-association (*P* = 6.3 x 10^- 6^). Twenty-nine of the 93 proteins from the MAM phase were also positively associated with WLZ in children with primary MAM, including those involved in musculoskeletal development [e.g., thrombospondin 3, collagen alpha-1(VI) chain, the aggrecan core protein present in articular cartilage, C-type mannose receptor 2 and leptin (regulator of bone metabolism as well as food intake and energy reserves) (*13*)], plus various facets of CNS development, vascular biology and cell adhesion (**table S8B**). In contrast, proteins negatively associated with WLZ were not significantly enriched in either treatment group of the current study (GSEA, *P* = 0.73). Although there was no overlap among the small number negatively WLZ-associated proteins between the MAM phases of the two studies (n=19 and 5), we noted a common functional theme related to various facets of immune activation (**table S8B**).

The second approach involved a correlated meta-analysis (CMA) (*14*) of plasma protein associations with WLZ obtained from the MAM-phase of the current study as well as the primary-MAM study. The strategy for CMA is illustrated in **Fig. 2A**. CMA reinforces *P*-values that support the same associations between separate studies (*14, 15*) (*see Methods*). To avoid an inflation of spurious *P*-values, CMA corrects for the degree of correlation between the plasma protein datasets by leveraging the basic biological assumption that most of the proteins will be under the null hypothesis of no association to any given outcome: i.e., only a relatively small proportion of the plasma proteome would be expected to have a true association to any particular outcome. To deal with the false-positives among the plasma proteins, the tetrachoric correlation coefficient is calculated, which is the maximum likelihood estimate of dichotomized *P*-values, so that true positive signals are not so influential in the correlation matrix estimates.

**Fig. 2.**
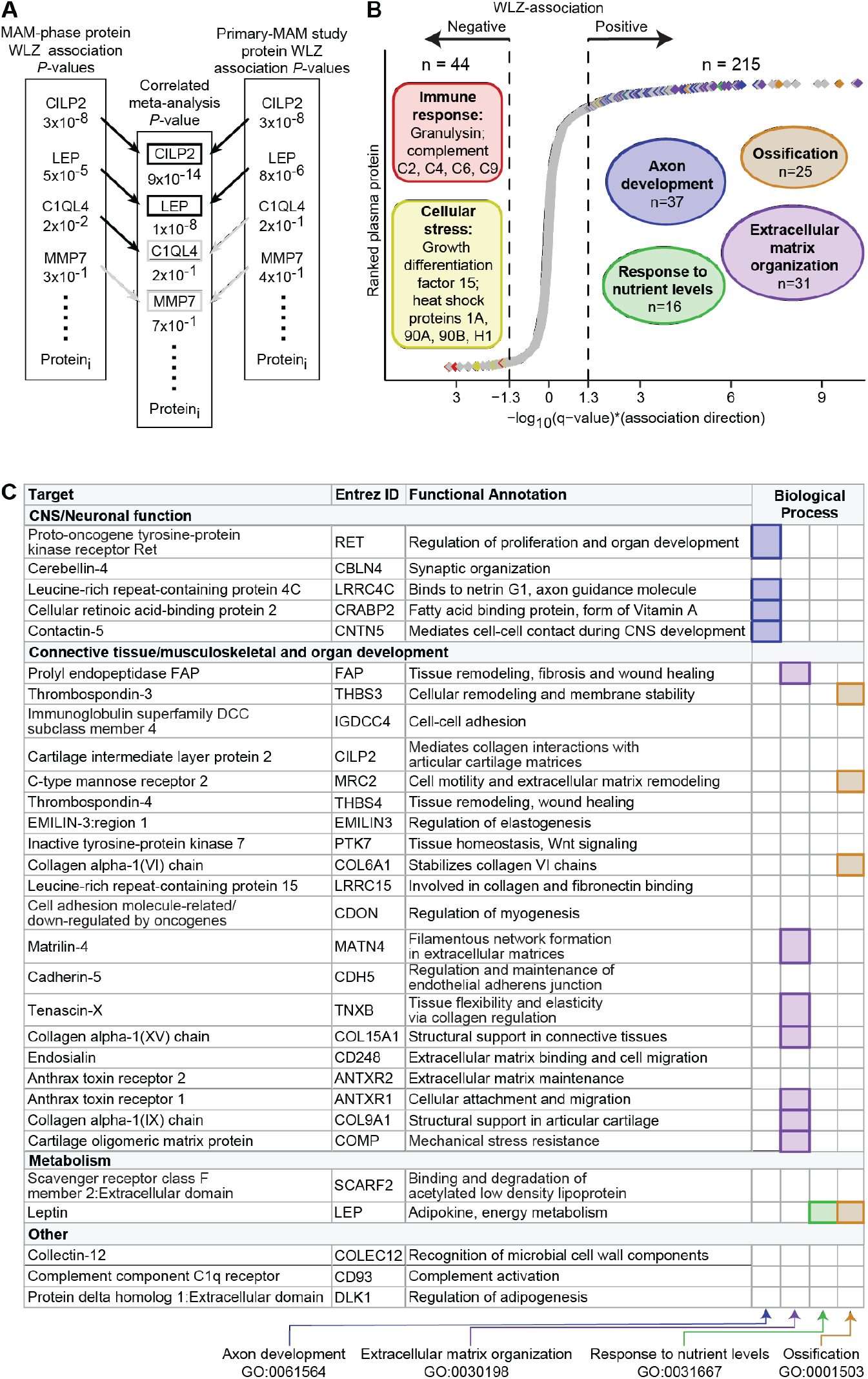
Correlated meta-analysis of plasma proteins. (**A**) Schematic of the Correlated Meta-Analysis (CMA). Black arrows and boxes indicate a protein with a WLZ-association *P*-value < 0.05, while grey arrows indicate a *P*-value > 0.05. (**B**) Plasma protein WLZ-associations obtained from CMA, with an over-representation analysis performed on proteins positively or negatively associated with WLZ. The colored circles indicate the Gene Ontology Biological Processes, with data points (proteins) colored by their membership in each GO Biological Process term. The number in each colored circle represents the number of proteins assigned to the GO BP. (**C**) The 30 most significantly WLZ-associated proteins from the CMA. The columns on the right summarize the assignments of proteins to GO Biological Processes.

CMA identified in 215 proteins positively associated with WLZ and 44 that were negatively associated (q-value < 0.05) (**Fig. 2B**; **table S9A**). We subsequently applied over-representation analysis of Gene Ontology (GO) ‘Biological Processes’ (*16*) to the group of proteins positively associated with WLZ, or separately to the group of proteins negatively associated with WLZ. The results revealed a statistically significant overrepresentation of positively WLZ-associated proteins in processes related to growth and development, including ossification (GO:0043931), extracellular matrix organization (GO:0030198), axon development (GO:0061564) and response to nutrients (GO:0007584), while the negatively WLZ-associated proteins were represented largely as components of immune and cellular stress responses (q-value < 0.05) (**table S9B,C; Fig. 2B**). The 30 most significantly WLZ-associated plasma proteins obtained from the CMA analysis are listed in **Fig. 2C** along with their GO-based functional annotations. All 30 proteins are positively associated with WLZ and include 20 involved in various aspects of connective tissue/musculoskeletal development (e.g., thrombospondin-3; prolyl endopeptidase FAP, tenascin-X), five regulators of neurodevelopment (e.g., the proto-oncogene tyrosine-protein kinase receptor Ret; cerebellin-4) and leptin, an adipokine involved in energy metabolism and satiety. The remaining 185 proteins encompass mediators of various facets of growth and development [e.g., insulin-like growth factor (IGF)-I; the liver-derived acid labile subunit of heterotrimeric complexes that contain IGF-1 and one of its IGF binding proteins; growth hormone receptor; lumican (collagen fibril assembly) (*17*); matrilin-2 and 3 (development of cartilage and bone) (*18*), BDNF/NT-3 growth factors receptor (synaptogenesis, neuronal development) (*19*); proopiomelanocortin (appetite, energy metabolism and stress responses) (*20*)]. As we had observed when our plasma proteomic analysis was restricted to the MAM treatment phase of the current study, the negatively WLZ associated proteins identified by CMA across both studies are broadly involved in inflammatory and stress responses (**Fig. 2B**).

We extended our CMA to identify MDCF-2- or RUSF-responsive changes in the levels of plasma proteins across the MAM treatment phase of the current study and in the primary MAM study. First, *P*-values obtained for the changes in their levels over the course of the intervention (**table S8A**, Eq. 12 and Eq. 13) were analyzed using CMA to gather consensus *P*-values of treatment association for each protein across both studies (**table S9D)**. The proteins were then ranked by the significance of their associations with MDCF-2 versus RUSF. Finally, we performed GSEA to test for enrichment of the 215 positively WLZ-associated proteins or 44 negatively WLZ-associated proteins in the MDCF-2 or RUSF arms. The results disclosed that proteins whose levels increased to a greater degree in response to MDCF-2 compared to RUSF treatment were significantly enriched in those that were positively associated with WLZ (q-value = 1.1×10^−6^), while the negatively WLZ associated proteins were not enriched in either treatment group (q-value = 0.94, **table S9E**). There were 67 leading edge proteins among the positively WLZ-associated proteins enriched in the MDCF-2 arm; they included thrombospondin-3, osteomodulin (bone mineralization), fibromodulin (extracellular matrix organization), matrilin 2, 3 and 4 (cartilage and bone development) (*21*), contactin 1, 4 and 5 (nervous system development) neuroplastin (synaptic plasticity and memory formation) (*22*), ROR1 (neurite growth) (*23*) and ROBO2 (axon guidance) (*24*) (**table S9E**).

Turning to the SAM treatment phase of the current study, we employed a linear modeling approach (see *Methods*) to identify 87 proteins that were significantly positively associated with WLZ and 12 proteins that were significantly negatively associated with WLZ (q-value < 0.05) (**table S8B**). Twenty-seven of the proteins that were positively associated with WLZ in the MAM phases of both studies (as defined by CMA) were also positively associated with the SAM phase (as determined by linear models); they included insulin-growth factor 1, growth hormone receptor, neutral ceramidase (sphingolipid digestion) (*25*) and tenascin C (cell adhesion and growth) (*26*) (**table S9F**). Of the 12 negatively WLZ-associated proteins from the SAM phase, none overlapped with our findings from CMA.

We concluded that (i) the set of WLZ-associated plasma proteins identified with CMA comprise a candidate panel of biomarkers for operationally defining recovery in the two groups of 12-18-month-old Bangladeshi children who had experienced two different ‘histories’ of presentation of MAM and (ii) compared to RUSF, MDCF-2 treatment is associated with a significant enrichment in levels of a group of 215 positively WLZ-associated proteins in children with MAM.

### Effects of treatment on the microbiome

Fecal samples were obtained from all participants when they presented with SAM and were enrolled in the study. Varying numbers of fecal samples were collected during their nutritional rehabilitation for SAM (prior to randomization), in accordance with the length of time required for each child to attain anthropometric recovery to a state of MAM. Samples were subsequently collected at fixed time points from each participant during the 3-month treatment with either MDCF-2 or RUSF and at the conclusion of the 1-month post-treatment follow-up (see **Fig. 1A** for a schematic of the sampling strategy). DNA was isolated from all fecal samples obtained from each trial participant (n=9-14; 760 in total) and used for qPCR assays of the abundances of 23 common protozoan, bacterial and viral enteropathogens, plus shotgun DNA sequencing to characterize the genomic features of bacterial strains (metagenome assembled genomes, MAGs).

#### Enteropathogen burden

Neither the abundance of any individual enteropathogen nor the total number of detectable enteropathogens were significantly related to the severity of SAM at enrollment, nor did they differ significantly between the Kurigram and Mirpur study sites (q > 0.05) (**table S10A**). The abundances of enterovirus, norovirus and enteropathogenic *E. coli* increased in children at both study sites during nutritional rehabilitation for SAM, while the abundance of *Campylobacter jejuni* decreased (paired Wilcox test, q < 0.05). Immediately prior to beginning treatment for MAM, we identified two enteropathogens with significant differences between the study sites: enterovirus was more abundant in children from Kurigram while Giardia was more abundant in children in Mirpur (Kruskal-Wallis test, q < 0.05; **table S10B**). Enteropathogen abundances were not significantly differentially impacted by MDCF-2 or RUSF treatment (q > 0.05; Kruskal-Wallis test; **table S10C,D)**.

#### Changes in MAG representation

**fig. S3** describes our pipeline for assembly of MAGs using shotgun sequencing data generated from each participant’s fecal samples (see *Methods*) (*9*). The resulting 754 high-quality bacterial MAGs (defined as ≥ 90% complete and ≤ 5% contaminated based on marker gene analysis; **table S11A**) were classified taxonomically with the Genome Taxonomy Database (GTDB) (*27*). The abundance of each MAG was determined by mapping data from each individual fecal sample to the set of high-quality, nonredundant MAGs collected across all trial participants (**table S11B**). Using this strategy, we assigned 73.6 ± 10.8% (mean ± sd) of all quality controlled, paired-end shotgun reads generated from all fecal DNA samples analyzed to these MAGs.

A principal component analysis (PCA) of MAG-based fecal microbiome configurations in fecal samples collected at enrollment, during acute rehabilitation, during MDCF-2/RUSF treatment, and at follow-up illustrated the similarities between children presenting with SAM at the two study sites (*P* > 0.05; PERMANOVA) with the most abundant MAGs assigned to the genera *Bifidobacterium, Escherichia, Prevotella*, and *Bacteroides/Phocaeicola* (**fig. S4**). At enrollment, the abundances of only 15 MAGs were significantly different between the study sites (q < 0.05; see Eq. 17 in *Methods* for the linear model).

The hospital-based rehabilitation protocol for SAM includes treatment with broad-spectrum oral and intravenous antibiotics, fortified milk and high energy density foods (**Fig. 1B**). This regimen significantly reduced the α-diversity (Shannon index) of their fecal microbiota (*P* = 8.99×10^−7^, see Eq. 18 in *Methods*; **table S11C**). Observations regarding β-diversity underscored the severity of this perturbation, as the Bray-Curtis dissimilarity of each child’s microbial community increased significantly after enrollment. The extent of β-diversity divergence from enrollment sample microbial community configurations was also related to the length of time each child required to complete acute rehabilitation (*P* < 2×10^−16^; Eq. 19) (**table S11C)**.

We used the PCA to characterize the dynamics of microbiome reconfiguration in trial participants. PERMANOVA within a space defined by the first three principal component (PC) projections, which together account for 35.3% of the overall variation in microbiome configuration (**table S12A)**, indicated significant shifts along the different treatment phases of the trial (*P* = 0.001). This analysis highlighted the prominent reconfiguration of the fecal microbiome associated with acute rehabilitation for SAM compared to enrollment (PERMANOVA, *P* = 0.001). Importantly, this process of reconfiguration continues but becomes less pronounced within the first two weeks of MDCF-2 or RUSF treatment (acute rehabilitation phase versus ‘baseline’ *P* = 0.068; acute rehabilitation versus MDCF-2/RUSF treatment *P* = 0.001) (**Fig. 3A)**.

**Fig. 3.**
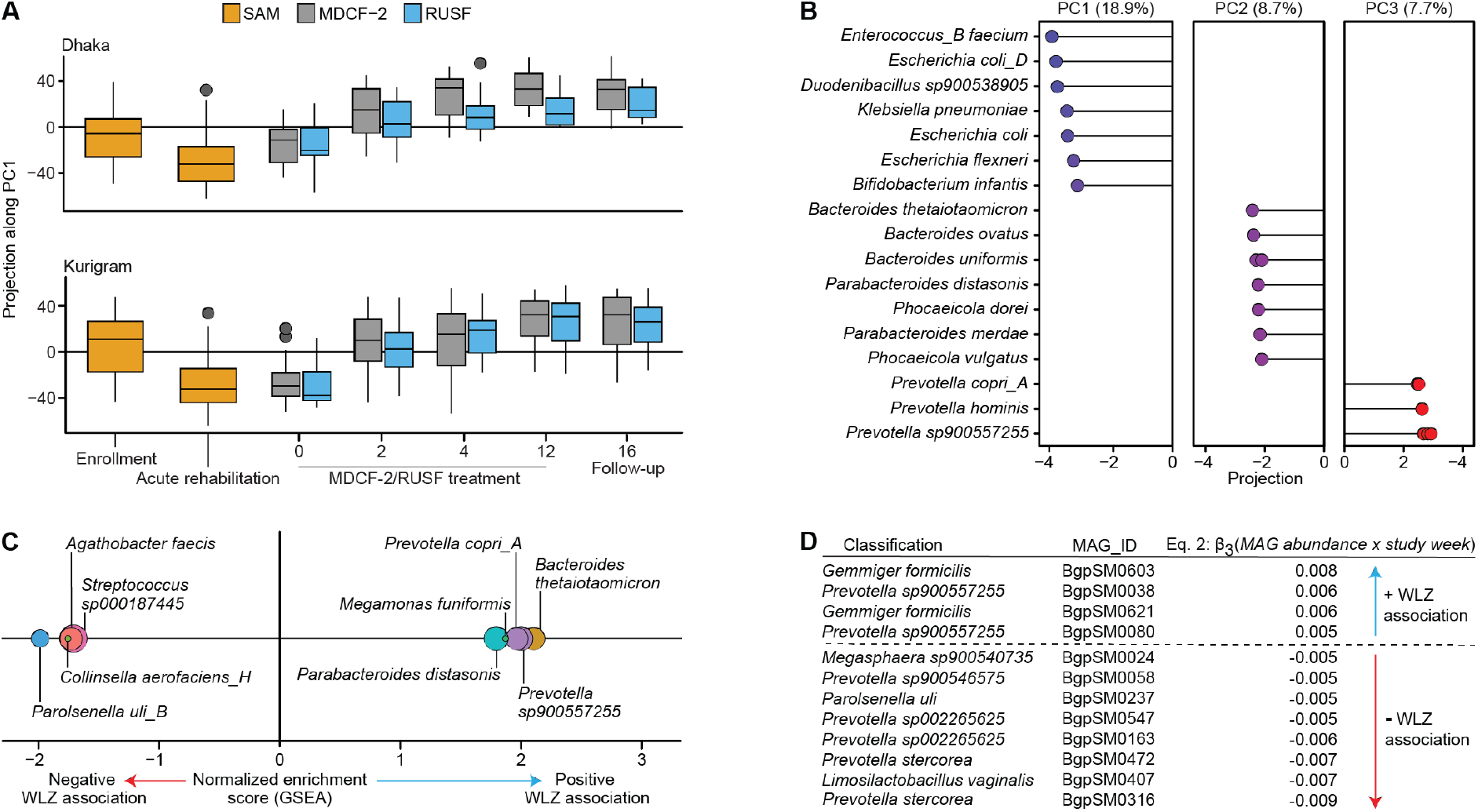
WLZ association of MAGs during acute rehabilitation for SAM and subsequent treatment for MAM with MDCF-2 or RUSF. Principal Component Analysis (PCA) was performed on variance-stabilized MAG abundances in fecal samples serially collected from participants through the course of the study. (**A**) Projections of fecal microbiome configurations along PC1 (representing 18.9% of total variance), separated by timepoint and study site during the SAM phase, and additionally by treatment group during the MAM phase and 1-month follow-up. Plots depict the median, first and third quartiles; whiskers extend to the largest value within 1.5 × the interquartile range. (**B**) Species-level taxonomic assignments of MAG ‘drivers’ of variance, defined as the top 1% of contributors to projection along each of the first three principal components. The percent variance explained by each principal component is indicated at top. (**C**) Results of enrichment analysis of MAGs, ranked by the strength and direction of their association with WLZ during acute rehabilitation for SAM. The horizontal axis indicates the normalized enrichment score, while the size of each point represents the number of MAGs assigned to each enriched species. (**D**) WLZ-associated MAGs identified in the MAM phase of the study.

We subsequently employed Singular Value Decomposition (SVD) to identify MAGs whose abundances were responsible for the microbiome reconfiguration observed in PC space. We defined “driver” MAGs for each PC as those comprising the top 1% of MAGs that contribute to the projection of samples along that PC. Drivers of PC1 included MAGs assigned to *Enterococcus faecium, Escherichia coli, Duodenibacillus sp900538905, Klebsiella pneumoniae, Escherichia flexneri*, and *Bifidobacterium longum* subsp. *infantis*. Projection along PC2 was driven by eight MAGs representing three species of *Bacteroides* (*B. thetaiotaomicron, B. ovatus* and *B. uniformis*), two species of *Parabacteroides* (*P. distasonis* and *P. merdae*) and two species of *Phocaeicola* (*P. dorei* and *P. vulgatus*). PC3 drivers included four MAGs annotated by GTDB as *Prevotella sp900557255*, three MAGs assigned to *Prevotella copri_A* and one to *Prevotella hominis* (**Fig. 3B; table S12B**). In aggregate, the MAG drivers of these three principal components can be viewed as biomarkers of the changes in microbiome structure that occur during acute rehabilitation for SAM, transition to MAM and initial response to MDCF-2 or RUSF treatment.

These findings raised the question of whether PC1 driver MAGs or MAGs contributing to additional PC projections are also associated with the change in WLZ we observed during treatment for SAM and/or MAM. Therefore, we analyzed the abundance of each MAG in all fecal samples collected from each study participant at both sites over the course of acute nutritional rehabilitation (*i*.*e*., prior to achieving a WLZ score above -3) (**table S13A,B**). To do so, we curated the MAG abundance dataset encompassing the entire trial by excluding low-abundance or infrequently detected MAGs (defined as those with abundances of <5 TPM in > 40% of samples), yielding a set of 613 MAGs of which 54% (n = 366) displayed significant changes in their abundances during acute rehabilitation of children at both study sites (q < 0.05; Eq. 20).

#### MAGs associated with WLZ during acute nutritional rehabilitation for SAM

At both sites, MAGs whose abundances *increased* the most during acute rehabilitation belonged to *Enterococcus faecium, Klebsiella pneumonia, Veillonella magna, Enterobacter hormaechei* and *Duodenibacillus* spp. The increased representation of these MAGs can be explained, at least in part, by an enrichment of genes in their genomes involved in resistance to the aminoglycoside and β-lactam antibiotics administered as part of the acute rehabilitation protocol [q < 0.05, gene set enrichment analysis (GSEA); **table S13C,D**]. We then used the linear model described in Eq. 1 to relate the rate of change of each of the 613 MAG abundances (**table S13A**) to the rate of change of WLZ between enrollment and the end of acute nutritional rehabilitation (the only timepoints in this phase of the study where anthropometry data were available for all participants at both study sites):

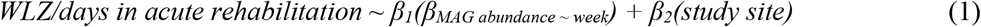

This analysis identified 120 MAGs with statistically significant associations with WLZ during the SAM phase (q < 0.05); 57 were positively associated and 63 were negatively associated (**table S14A**). GSEA performed on these MAGs ranked according to the strength and direction of their association with WLZ revealed that those positively associated with WLZ during nutritional rehabilitation for SAM were enriched for two members of the *P. copri* lineage (*Prevotella copri_A* and *Prevotella sp900557255*, (n=12 and n=11 MAGs, respectively), plus *Parabacteroides distasonis* (n=12 MAGs), *Bacteroides thetaiotaomicron* (n=11 MAGs) and *Megamonis funiformis* (n=5 MAGs) (q-value < 0.05, **Fig. 3C**).

MAGs negatively associated with WLZ in this phase were enriched for *Agathobacter faecis* (n=11), *Parolsenella uli* (n=8) and *Collinsella aerofaciens* (n=5) (**table S14B**). Although these 120 MAGs were associated with WLZ in this phase, none of their abundances at enrollment individually predicted the subsequent rate of anthropometric recovery of children from SAM [q > 0.05 (dream); see Eq. 21 in *Methods*].

#### MAGs associated with WLZ during the MAM phase

Fecal microbiome samples and anthropometric data were collected at regular intervals during treatment for MAM. Analyses using the mixed effects linear model shown in Eq. 2 yielded 12 MAGs whose abundances over time were significantly associated with WLZ (q < 0.1) (**Fig. 3D**; **table S14C**).

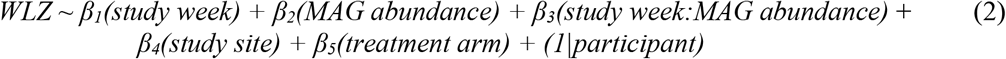

Four of these MAGs displayed positive relationships with WLZ; two were assigned to *Prevotella sp900557255*, including BgpSM0038 which had a positive relationship with WLZ in both the MAM and SAM phases, and BgpSM0080 which was positively associated with WLZ only in the MAM phase. The other two MAGs were classified as *Gemmiger formicilis* (BgpSM0603, BgpSM0621); their significant positive association with WLZ was limited to the MAM phase. Interestingly, two of the 14 *G. formicilis* MAGs identified in our study of comparably aged Bangladeshi children with primary MAM were also positively associated with WLZ (*9*). Furthermore, we observed a significant positive association between the abundances of *Prevotella sp900557255* BgpSM0080 and *G. formicilis* BgpSM0621 during treatment for MAM (q=0.01; see Eq. 22 in *Methods*), suggesting potential ecological relationships between these organisms (**table S14D**). As in the previous MDCF-2 trial, no individual MAG had a statistically significant different change in its abundance over the 3-month intervention when comparing the MDCF-2 and RUSF treatment groups (q>0.1; **table S14E**).

We performed a marker gene-based phylogenetic analysis to more precisely contextualize the relationship between these two *Prevotella sp900557255* MAGs that either exhibited a positive relationship with WLZ in both the SAM and MAM phases (BgpSM0038) or only during the MAM phase (BgpSM0080). The analysis included (i) all *Prevotella* MAGs identified in this study regardless of their WLZ association, (ii) the 11 *P. copri* MAGs identified in our prior primary MAM study, including the two that were significantly positively associated with WLZ (Bg0018 and Bg0019) (*9*), (iii) *P. copri* strains (Bg131, BgF5_2 and BgD5_2) cultured from Bangladeshi children living in Mirpur that we have shown to be key mediators of MDCF-2 glycan metabolism and key effectors of fatty acid and amino acid metabolism in the intestinal epithelium of gnotobiotic mice (*10*), and (iv) publicly-available reference genomes representing a diversity of clade-level *P. copri* designations and related *Prevotella* species (Publicly available cultured *P. copri* isolates and MAGs have been taxonomically assigned clades designated A-D) (*28*). The results revealed that the positively WLZ-associated *Prevotella sp900557255* MAGs identified in the current study belong to a branch of *P. copri* clade ‘A’ that also contains the WLZ-associated *P. copri* MAGs identified in the prior primary MAM RCT (**Fig. 4A**).

**Fig. 4.**
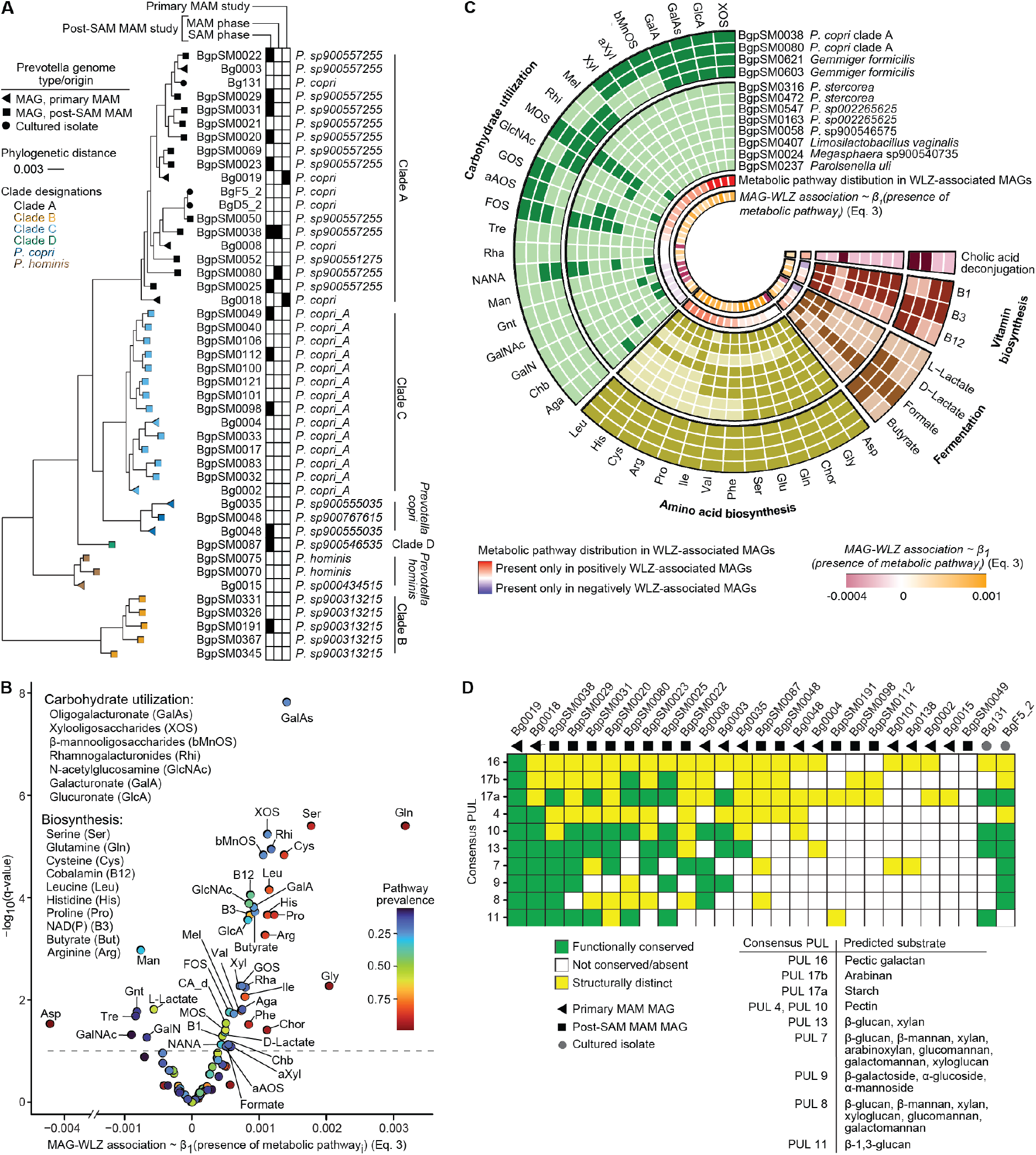
Phylogenetic and functional characteristics of MAGs associated with WLZ in the SAM and MAM phases of the trial. (**A**) Phylogenetic analysis of the relationships of WLZ-associated *P. copri* and other *P. copri* MAGs from the current study, *P. copri* MAGs identified in our prior study of similarly aged (12-18-month-old) Bangladeshi children with primary MAM, and selected *P. copri* isolates cultured from the study population. The black boxes in the grid denote MAGs that displayed significant WLZ-associations during the indicated phases of this or the prior MDCF-2 study. Tree tip colors on the left side of the panel refer to the clade assignment of each MAG while the shape of the tips (square, triangle) indicate the origin of each MAG. (**B**) Relationship between the presence of mcSEED metabolic pathways and the WLZ associations of MAGs possessing each pathway. Points are colored based on the prevalence of each metabolic pathway across the set of 613 MAGs. Pathways whose presence or absence were significantly associated (q < 0.001) with the MAG WLZ-association coefficient (β3 in Eq. 2) are listed on the right in order of decreasing association strength. The dashed line represents the significance threshold (q < 0.05). (**C**) The presence or absence of mcSEED metabolic pathways in MAGs positively associated with WLZ during the MAM phase (outer four rings) compared to negatively WLZ-associated MAGs (next eight inner rings). The second innermost ring denotes the prevalence of each metabolic pathway in positively versus negatively WLZ-associated MAGs. The innermost ring indicates the strength of the association of each predicted phenotype with the WLZ association of each MAG. Only pathways with a significant *β*_1_ coefficient are included (q < 0.1). (**D**) PUL conservation in WLZ-associated *P. copri* MAGs identified in the current and prior MDCF-2 trial compared to other *P. copri* MAGs and isolates cultured from the study population. The color-coded matrix in the center indicates the extent of conservation each PUL in each MAG relative to Bg0019 (the MAG with the strongest WLZ association in the fecal microbiomes of participants in the primary MAM trial). The known or predicted polysaccharide substrates of these PULs are described (see **table 16B** for details of PUL annotation and numbering convention).

A similar phylogenetic analysis of the five *G. formicilis* MAGs including the two that were positively associated with WLZ, plus the 17 *Gemmiger* MAGs from our previous study of children with primary MAM revealed that, unlike our observations for *P. copri*, WLZ-associated *G. formicilis* MAGs did not cluster together within the marker-gene based phylogeny or when genome relatedness was judged by average nucleotide identity (**fig. S5**).

In summary, our characterization of the trajectories and MAG drivers of fecal microbial community dynamics using diversity metrics and PCA disclosed a significant impact of antibiotic treatment and nutritional resuscitation on microbiome configuration during recovery from SAM that extended into the early MAM treatment phase. Neither the position of each sample within a PCA space defined by microbiota configuration (q > 0.05, Eq. 24) nor the abundance of any single MAG in fecal samples collected at enrollment or baseline (q > 0.05, Eq. 25) was predictive of the rate of change of WLZ during MDCF-2 or RUSF treatment for MAM.

There was a remarkable consistency in the abundances of *Prevotella* MAGs between the two sites just prior to initiation of treatment with MDCF-2 or RUSF. The 63 MAGs belonging to *Prevotella* were detected in 99.8% of samples collected at baseline. All samples collected at baseline from all participants contained the two positively WLZ-associated *P. copri* MAGs. Moreover, none of the 33 *P. copri* or five *G. formicilis* MAGs, including those associated with WLZ, were differentially abundant between study sites (Eq. 17; **table S13B**; q < 0.05).

#### Functional features of WLZ-correlated MAGs

We employed a ‘metabolic subsystems’ approach adapted from the SEED genome annotation platform to identify encoded functions that distinguished WLZ-associated MAGs from those that were not significantly associated with ponderal growth (*29*). Genes were first aligned to a reference collection of 2,856 human gut bacterial genomes that had undergone *in silico* reconstructions of metabolic pathways involved in major nutrient biosynthetic and degradative activities within a microbial community-centered implementation of SEED (mcSEED) (*30*). Putative functions were assigned to 194,453 proteins in the 613 MAGs comprising our abundance-filtered dataset and using procedures described in *Methods, in silico* predictions were made regarding which of 106 metabolic pathways were present or absent in each MAG (**table S15A-C**).

We then used the linear model shown in Eq. 3 below to relate the presence or absence of each metabolic pathway in each of the 613 MAGs to their WLZ association coefficients (*β*_3_ in Eq. 2) during the MAM phase of the study:

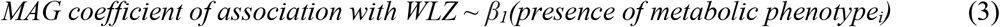

The results disclosed 38 pathways whose presence in MAGs was predictive of a positive WLZ association and seven whose presence was predictive of negative association (q < 0.1, **Fig. 4B; table S15D**). Comparing the representation of these metabolic pathways between the four positively WLZ-associated MAGs in the MAM phase of the study and the eight MAGs that were negatively associated (**Fig. 4C**) revealed that the prevalence of carbohydrate utilization pathways was significantly greater in the four positively WLZ-associated MAGs (Wilcoxon rank sum test, *P* = 0.006). Both *P. copri* BgpSM0038 and BgpSM0080 contain pathways for metabolizing carbohydrates present in MDCF-2; i.e., pathways involved in the utilization of β-mannooligosaccharides, α-xylosides, galacturonate, oligogalacturonate, glucuronate, galactooligosaccharides, xylooligosaccharides and xylose. Several of these pathways (xylooligosaccharide, glucuronate, oligogalacturonate, galacturonate and maltooligosaccharide utilization) were also represented in the positively WLZ-associated *G. formicilis* MAGs while none were represented in the eight MAGs identified as negatively associated with WLZ (**fig. S5B; table S15C**).

The number and type of mcSEED carbohydrate utilization pathways were similar across the 33 *P. copri* MAGs we identified in trial participants and did not distinguish the *P. copri* MAGs that were or were not significantly associated with WLZ (Fisher’s exact test, *P* = 0.996, **fig. S6**). Another way of comparing the carbohydrate utilization machinery in different *P. copri* MAGs is to examine their repertoires of polysaccharide utilization loci (PULs). Our bioinformatic approach to identifying PULs integrates (i) searches for genes encoding SusC and SusD homologs (PUL marker genes that mediate active uptake of poly/oligosaccharides across the outer membrane of related saccharolytic *Bacteroidaceae* species) (*31*) plus genes encoding CAZymes with (ii) synteny information to identify gene clusters (loci) predicted to coordinate degradation of specific polysaccharides (*32*). The putative substrates of each PUL can be inferred from the known or predicted specificities of its component CAZymes. As noted in the *Introduction*, the PUL repertoire of the two *P. copri* MAGs that were significantly positively associated with WLZ in the primary MAM study distinguished them from the nine other *P. copri* MAGs that did not display this association with ponderal growth (*9*). Therefore, we compared the PUL repertoires of (i) the 14 *P. copri* MAG identified in the current study as being WLZ-associated in the MAM phase, SAM phase or both, (ii) the 11 *P. copri* MAGs characterized in our primary MAM study, and (iii) the cultured Bangladeshi *P. copri* strains whose *in vitro* growth we had quantified in the presence of purified polysaccharides representing various MDCF-2 glycan structures (*9*).

Remarkably, among the *P. copri* MAGs identified in the current study, the MAG with the strongest WLZ association during the MAM treatment phase (BgpSM0038) had the most homologous PUL profile to the two WLZ-associated *P. copri* MAGs (Bg0018 and Bg0019) identified in the primary MAM study (**Fig. 4D, table S16A**). We used the CAZymes encoded by the PULs conserved between these three MAGs to infer that their polysaccharide targets included β-glucan, β-mannan, xylan, glucomannan, galactomannan, β-galactosides, α-mannosides, α-glucoside, arabinoxylan, xyloglucan, starch and arabinogalactan. Notably, the conserved PUL 7 homologs in Bg0019, Bg0018 and BgpSM0038 encode a bifunctional GH26|GH5_4 CAZyme predicted to cleave a diverse set of dietary glycans, including β-glucan, β-mannan, xylan, arabinoxylan, glucomannan, galactomannan and xyloglucan, which are unique to and/or more abundant components of MDCF-2 compared to RUSF (*9*). Although *P. copri* BgpSM0080, the other positively WLZ-associated MAG from the MAM treatment phase of the current study, lacks a conserved PUL 7 homolog, its genome encodes three PULs with specificities for a subset of PUL 7 targets, including β-glucan, pectin, starch, and arabinogalactan (**Fig. 4D**).

Finally, we examined whether the positively WLZ-associated MAGs assigned to *P. copri* and *G. formicilis* could share ‘complementary’ or ‘cooperative’ roles in the metabolism of MDCF-2 glycans. Two possible examples are (i) complementary metabolism of pectins such as rhamnogalacturonan and homogalacturonan and (ii) cooperative metabolism of xylan and arabinoxylan. Mass spectrometry has revealed key differences in the abundances and structures of members of these classes of glycans in MDCF-2 and RUSF (*9*). Although the two WLZ-associated *P. copri* MAGs encode CAZymes that have a broad array of substrate specificities, most are predicted to target plant mannans and xylans. *G. formicilis* BgpSM0603 and BgpSM0621 contain a smaller number of xylan-targeting CAZymes; most of their CAZymes are predicted to predominantly target pectins (**table S16B**). These observations suggest that these organisms have complementary roles; each organism has the potential to degrade distinct components of MDCF-2. In contrast to their differing roles in pectin degradation, *P. copri* and *G. formicilis* may have a cooperative role in degrading xylans. *In vitro*, Bangladeshi *P. copri* isolates similar to BgpSM0038 and BgpSM0080 (**Fig. 4D**) are capable of utilizing xylans through the activities of their PULs and component CAZymes. Efficient degradation of these xylans requires both endo- and exo-acting CAZymes, which are numerous in the genomes of WLZ-associated *P. copri* MAGs. However, the xylan or xylooligosaccharide-targeting CAZymes of *G. formicilis* MAGs are exclusively exo-acting (**table S17**). Thus, it is possible that *P. copri* performs primary (endo) degradative activities and both *P. copri* and *G. formicilis* participate in secondary degradation of xylooligosaccharides. Direct proof of the capacity of the strains represented by these MAGs to collaborate and/or cooperate in the metabolism of MDCF-2 glycans will require culturing the corresponding *G. formicilis* strains and performing *in vitro* assays of their growth in defined medium supplemented with various glycan structures, as well as *in vivo* studies of their contributions to MDCF-2 metabolism using preclinical models (e.g., in gnotobiotic mice).

## DISCUSSION

We have tested the ability of a microbiome-directed therapeutic food, MDCF-2, to improve the ponderal growth rates (β-WLZ) of acutely malnourished 12-18-month-old Bangladeshi children living in both urban and rural settings who had completed a hospital-based rehabilitation program for severe acute malnutrition but were still wasted (i.e., had moderate acute malnutrition). In this randomized controlled trial, treatment of children with MDCF-2 with post-SAM MAM produced a superior rate of weight gain than treatment with a calorically more dense, standard ready-to-use supplementary food (RUSF). This result is similar to what we had previously reported in a comparably designed study of 12-18-month-old Bangladeshi children with primary MAM (i.e., those without an antecedent history of SAM) (*7*), despite children with post-SAM MAM having significantly worse anthropometric measures of ponderal and linear growth. In both studies, though wasting was frequently accompanied by stunting, MDCF-2 did not produce a statistically significant effect on LAZ compared to RUSF during the 3-month intervention period. However, MDCF-2 led to a significant improvement in linear growth trajectories of children with primary MAM in the subsequent 2-year period of follow-up (*8*); this effect was foreshadowed by increased levels of plasma proteomic biomarkers and mediators of musculoskeletal development at the end of the treatment phase. Longer periods of intervention and/or more extended post-treatment follow-up will be needed to determine whether stunting, and/or neurodevelopmental abnormalities, are ameliorated by MDCF-2 in children with post-SAM MAM.

This trial of MDCF-2 in 12-18-month-old children with post-SAM MAM and the previous trial involving those with primary MAM provide evidence that *P. copri* is both (i) a target of MDCF-2 and (ii) linked to the ponderal growth responses in both patient cohorts. The underlying strain specificity is remarkable; the two WLZ-associated *P. copri* MAGs in this study and the two identified in the previous study are phylogenetically closely related members of *P. copri* clade A; they are distinguishable from the non-WLZ associated clade A *P. copri* MAGs present in the gut (fecal) microbial communities of study participants based on their content of PULs known or predicted to target glycans present in MDCF-2. These observations, combined with the results of (i) our growth assays of cultured Bangladeshi *P. copri* strains, whose repertoires of PULs correspond to those in the MAGs, in defined media supplemented with glycan structures present in MDCF-2 (*9*) and (ii) our ‘reverse translation’ studies in gnotobiotic mice demonstrating the importance of cultured Bangladeshi *P. copri* strains embodying these genomic features in mediating MDCF-2 glycan utilization, energy metabolism in gut epithelial cells, and weight gain (*10*), provide an expanding body of evidence that *P. copri* functions at the intersection between MDCF-2 metabolism and rescue of growth faltering in Bangladeshi children with MAM. The genomic features that distinguish these growth-promoting strains (MAGs) (notably PULs) provide a way for characterizing their distribution in human populations, including women and children living in different low- and middle-income countries where the burden of undernutrition is great. Their PULs also point to the bioactive glycan structures present in MDCF-2 – structures that if captured from sustainable and scalable sources in an affordable manner could form a future prebiotic therapeutic for undernutrition.

A major challenge is to define the microbial metabolites that mediate host responses to MDCF-2 treatment. For example, we do not know whether these microbial metabolites are the products of MDCF-2 glycan metabolism per se (and/or other components of MDCF-2), or whether they are the products of metabolic pathways whose activities in *P. copri* or other community members are affected by the metabolism of MCDF-2 glycans. Obtaining answers should be facilitated by re-enacting the process of gut microbial community assembly that occurs in postnatal development in gnotobiotic mouse experiments that involve maternal (dam)-to-pup transmission of cultured, genome-sequenced bacterial strains corresponding to the age- and growth-discriminatory MAGs we identified in the study population. These types of ‘reverse translation’ experiments can be designed with multiple arms where none, one or more of the cultured strains are ‘left out’ of the defined communities so that their contributions to MDCF-2 induced microbial and host metabolic and growth responses can be determined. Given the large number of combinatorial possibilities, these envisioned manipulations of community composition underscore the general need of the field of human microbiome research to develop scalable unbiased methods for determining which members of a microbiota influence specific metabolic and physiologic phenotypes (*33*).

An underlying principle of this longitudinal study was that each individual served as her/his own control; i.e., measurements of anthropometric values, MAG representation, and the plasma proteome prior to, during and at the conclusion of an intervention are used to define individual responses to treatment; the level of similarity in these responses between members of a given treatment group and differences from responses exhibited by members of another treatment group can then be ascertained. The need to make comparisons within and across individuals provided a justification for first performing a *de novo* assembly of high-quality MAGs present in the microbiome of each individual and then finding corresponding MAGs across all individuals and treatment groups before analyzing MAG associations with WLZ responses. In the current study, anthropometry alone guided the timepoint selected for beginning treatment of MAM with MDCF-2 or RUSF. The standard protocol used to treat SAM in these Bangladeshi children involved stabilization, plus a short course of oral and intravenous antibiotics along with administration of fortified infant formula, followed by a calorically rich rice and lentil-based nutritional supplement. This protocol, although supporting anthropometric recovery from SAM, produced a pronounced reduction in bacterial (MAG) diversity and was associated with a period of substantial microbiome reconfiguration that had not stabilized prior to initiation of MDCF-2 or RUSF treatment. As is customary, we defined the transition between SAM and MAM based on anthropometry alone. This sustained period of microbiome reconfiguration raises the question of whether additional measurements beyond anthropometry, such as the plasma proteome, should be used to define the ‘beginning’ of the MAM phase and what is designated as the ‘baseline’ state of the microbiome and host prior to initiation of MDCF-2 and RUSF treatment.

Our study had several limitations, including the exclusion of 50 of the original enrollees from the multi-omics analysis, with a resulting reduction in statistical power, due to a severe flood which affected one of the study sites. In addition, the study design included hospitalization and a course of oral and parenteral antibiotics which confound attempts to define the true ‘baseline’ metagenomic and plasma proteomic states of children with post-SAM MAM. However, a more accurate determination of how anthropometric, metagenomic and plasma proteomic features of children with post-SAM MAM differ from those with primary MAM should be forthcoming from an upcoming randomized controlled trial (*34*). This trial will enroll children who present with uncomplicated SAM and not involve hospitalization or extensive antibiotic therapy. Another limitation relates to technical challenges with inter-run normalization using different versions of the SOMAscan platform that prevented direct comparisons of protein levels between the two studies at a given timepoint (e.g., just prior to initiation of intervention). Therefore, our analytical strategy was based on characterizing changes in the levels of proteins from the beginning to the end of treatment in each study. Using a comparative meta-analysis of the levels of 4,520 plasma proteins during the course of the treatment phase of this study and in our previously reported primary MAM trial, we identified 259 shared biomarkers of recovery from MAM. Among these, proteins that were positively associated with WLZ are enriched in themes (GO Biological Processes) related to musculoskeletal development, neurodevelopment and energy metabolism, while negatively WLZ-associated proteins were enriched in immune and stress response-related themes. We also demonstrated that the positively WLZ-associated proteins are increased in the plasma of MDCF-2 treated children compared to those treated with RUSF in both studies.

These 259 plasma proteins represent biomarkers to evaluate in future studies involving MDCFs in children with acute malnutrition, living in different geographic locales, and having varying degrees of disease severity, as well as in studies involving other therapeutic agents. Goals will be to determine the utility of these biomarkers for (i) stratifying children prior to treatment by providing a more comprehensive definition of their initial physiologic state(s), (ii) judging the efficacy, generalizability and durability of MDCF treatment and (iii) predicting treatment outcomes, including those related to ponderal growth and perhaps, in longer post-treatment follow-up studies, linear growth as well as neurodevelopment.

## Methods

### Study design and execution

The human study entitled ‘Community-based Clinical Trial With Microbiota Directed Complementary Foods (MDCFs) Made of Locally Available Food Ingredients for the Management of Children With Post-severe Acute Malnutrition Moderate Acute Malnutrition (Post-SAM MAM)’, was approved by the Ethical Review Committee at the icddr,b (Protocol PR-18079; ClinicalTrials.gov identifier: NCT04015986). Informed consent was obtained for all participants. The objective of the study was to determine whether twice daily, controlled administration of a locally produced, microbiota-directed complementary food (MDCF-2) for 3 months to children with post-SAM MAM in urban and rural locations led to superior improvements in WLZ, microbiota repair, and in the levels of key plasma biomarkers/mediators of healthy growth compared to a standard rice/lentil-based ready-to-use supplementary food (RUSF) formulation.

Children aged 12- to 18-months with WLZ < -3 who did not exhibit bilateral pedal edema were enrolled. Written consent was obtained from the children’s parents. Children with congenital abnormalities, soy, peanut, or milk protein allergies, tuberculosis, or severe anemia (8 mg/dL) were excluded from the study. Children with SAM were identified at Dhaka Hospital, RADDA-MCH Clinic in Mirpur, the Special Nutrition Unit of Terre des Hommes in Kurigram, and through community health screenings conducted in Mirpur and in different areas of the Kurigram district. These facilities admitted children who required stabilization, providing them with standard care during the acute phase of treatment for SAM. Acute nutritional rehabilitation consisted of two-phases: (i) a standardized protocol including rehydration, administration of antibiotics, micronutrient supplementation, plus timely management of any concomitant complications (*12*), and (ii) a standard care dietary protocol based on F-75/F-100 therapeutic milk followed by high energy local diets, including Halwa and Khichuri, until children attained a WLZ between -3 and -2. Breastfeeding was also encouraged throughout the study.

After acute nutritional rehabilitation, children entered the 3-month MAM intervention phase where they were randomized to the MDCF-2 or RUSF treatment groups; allocation was determined through sealed, non-transparent envelopes that contained the assigned treatment code. The study gathered socio-demographic information, anthropometric measurements, data on food diversity, as well as samples of blood, feces and urine. Food Frequency Questionnaires were used to collect weekly qualitative dietary intake data over a 24-hour period.

Both MDCF-2 and RUSF were produced locally at the study centers in Mirpur and Kurigram using ingredients and procedures described in previous publications (*7, 35*). Food preparation was conducted daily as a precautionary measure to prevent contamination or nutrient depletion due to spoilage. Raw ingredients were sourced from local markets. Preparation, dispensing, and feeding of MDCF-2 and RUSF took place on the same day.

Treatment with MDCF-2 or RUSF was designed to provide 20-25% of the daily energy requirements of participants. Mothers or primary caregivers were encouraged to maintain each child’s usual dietary and breastfeeding routines, and were asked to bring their children to established nutrition centers at the study sites twice daily during the first month and once daily during the second month (one treatment was administered at home during this second month). During the third month, the children were fed the supplement twice daily at home (**Fig. 1A**). Mothers were provided with explicit instructions to refrain from providing any food or breast milk to their children for two hours preceding the designated mealtime. Each participant was allocated two 25 g servings of MDCF-2 or RUSF per day. Children were spoon-fed the pre-weighed diets until they refused to eat. After a brief interval of two minutes, they resumed the feeding session, continuing until the child refused. Following a second two-minute interval, the diet was offered for a third time until the children refused again. At the point of this third refusal, the feeding session was considered terminated, and any unconsumed supplement was weighed.

To ensure sample integrity for subsequent DNA analyses, fecal biospecimens were collected within 20 minutes of their production and immediately transferred to liquid nitrogen-charged vapor shippers for transport to a -80 °C freezer at the nearest study center. Coded biospecimens (plasma and feces) were shipped to Washington University on dry ice where they were stored at -80 °C in a dedicated biorepository with approval from the Washington University Human Research Protection Office.

### Anthropometry

#### Enrollment and baseline analyses

Comparisons of demographic, anthropometric, and environmental features at enrollment (presentation with SAM) or baseline (immediately prior to treatment with MDCF-2 or RUSF) between children receiving MDCF-2 or RUSF were performed as previously described (*7*). Briefly, we employed two-sided unpaired t-tests for numerical data or Fisher’s exact tests for categorical data. For all time course analyses of anthropometric measurements, the first day of the corresponding intervention (acute rehabilitation or MDCF-2/RUSF treatment) was used as the baseline measurement. Comparisons of enrollment (SAM) or baseline (post-SAM MAM) anthropometric measures were performed using unpaired t-tests and are reported between children at each study site or between children at each site who subsequently received MDCF-2 or RUSF (**table S3**). To compare anthropometric measurements at baseline between the current study and primary-MAM study (*7*), we applied the following linear model:

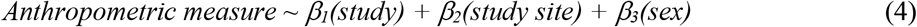

#### Analysis of the anthropometric effect of flooding in Kurigram

To test the effects of flooding during the course of the MAM phase intervention, we applied the following linear mixed-effects model to anthropometry data collected from trial participants who had otherwise completed the study per-protocol:

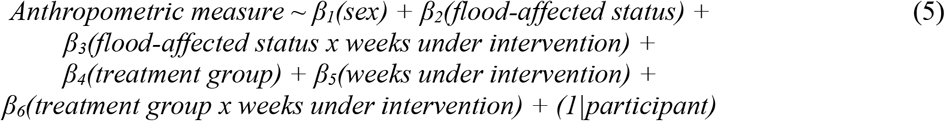

#### Analysis of anthropometric responses to MDCF-2 or RUSF

Trial participants who did not complete the nutritional intervention per protocol were excluded from primary analyses of anthropometric responses (**table S1A, fig. S2**). Ponderal growth rate within each treatment group was calculated using a mixed-effects linear model that related each anthropometric measure to weeks after initiating the corresponding MDCF-2 or RUSF intervention, controlling for sex, study site, weeks under intervention and the interaction between study site and weeks under intervention, and a random intercept for each participant ID. We employed the following model:

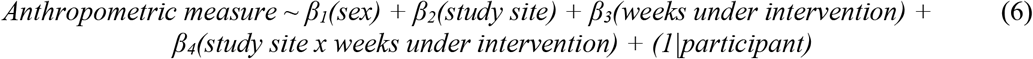

Note that all linear mixed effects models described in this report were performed using the R packages lme4 (v1.1-27.1) (*36*) and lmerTest, (v3.1-3) (*37*). Mean rates of growth for children receiving either MDCF-2 or RUSF are reported in **table S6** and represent the weekly increase in WLZ, WAZ, MUAC or LAZ within a given treatment arm. An analysis of differential effect of MDCF-2 versus RUSF treatment on growth rates was performed using a similar mixed effects linear model but with the addition of terms describing treatment group and the interaction between treatment group and weeks under intervention:

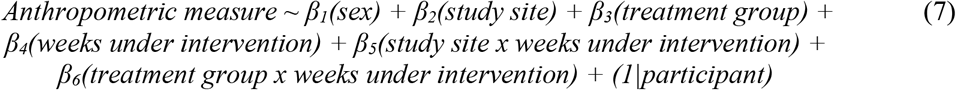

The differential rate of change in WLZ, WAZ, MUAC or LAZ as a function of treatment arm (MDCF-2 versus RUSF) is reported in **table S6**.

For analyses performed within or between each treatment group, linear mixed effects models were run on datasets that included (i) only anthropometric measurements from the 3-month-long MAM treatment phase, or (ii) anthropometric measurements collected during the MAM treatment phase plus the additional 1-month follow-up timepoint. An additional ‘Intent-to-Treat’ analysis was conducted using the models described in Eq. 6 and Eq. 7 but without exclusion of participants due to trial protocol noncompliance, or the effect of flooding. Results of these analyses are reported in **table S6A,D**.

### Food frequency questionnaire and comorbidity analysis

Food frequency questionnaire responses were collected at enrollment, during the acute rehabilitation phase, and at week 0, 1, 2, 4, 8 and 12 of MDCF-2 and RUSF treatment. To search for differential responses between the MDCF-2 and RUSF treatment groups, we applied generalized linear models to each question. These models employed a Poisson regression for questions that provided count answers, and binomial regression for questions with yes or no responses:

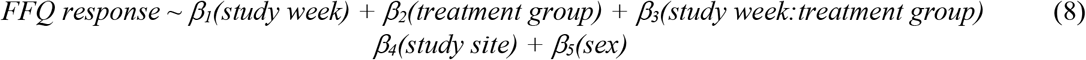

To search for differential responses by treatment arm at enrollment, at baseline and at the end of treatment, we applied generalized linear models with the same Poisson and binomial regression selection strategy employed in Eq. 8:

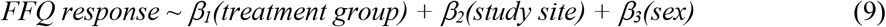

To identify if the occurrence of comorbidities were associated with ponderal growth responses, the total number of days with a reported comorbidity (cough, fever, runny nose and diarrhea) throughout the MAM phase for each participant was regressed against their rate of change of WLZ using a linear model:

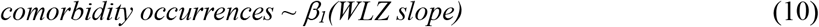

A binomial generalized linear model was used to test if the occurrences of comorbidities (recorded daily) were significantly different over time between treatment groups:

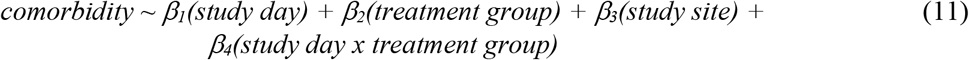

### Aptamer-based analysis of the plasma proteome

*See Supplemental Methods for sample collection and plasma protein quantification*.

#### Linear models

The change in levels of plasma proteins between baseline and end of MDCF-2 and RUSF treatment were identified using the following linear model:

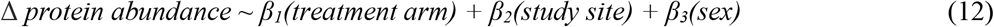

A similar model was applied to find treatment specific differences of plasma protein levels in the primary MAM study:

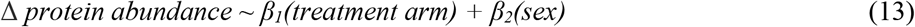

The rate of change of WLZ was calculated for each participant_i_ during the SAM phase (from enrollment to baseline) and the MAM-phase (rate of change of WLZ per week), while the changes in plasma protein_j_ levels for each participant_i_ were obtained by the change between the beginning and end of the SAM phase and the beginning and end of the MAM phase. The association between the level of each plasma protein and WLZ was determined using the two linear models listed below:

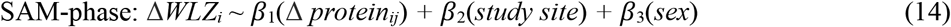

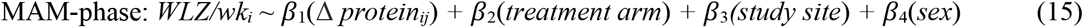

The statistical significance (*P-*value) of the association of each plasma protein with WLZ was determined using ANOVA, with false discovery rate (FDR) controlled using a Benjamini-Hochberg correction (**table S8B**).

#### Correlated meta-analysis (CMA) of the plasma proteome

The suitability of our plasma proteomic datasets for CMA was first established by testing for structural bias, which could lead to an inflation of false positive or negative discoveries under our modeling strategy. To do so, the metadata (WLZ/week, treatment group, study site and sex) for each protein dataset was first shuffled, after which relationships with the rate of change of WLZ were evaluated using Eq. 15. The *P*-value distribution of the *β*_1_(Δ *protein*_*ij*_) coefficients was then tested for excess false discoveries using the inflation factor λ (*38*). This evaluation process was repeated 10,000 times to test whether the median λ was < 1.1, which is an accepted threshold for detecting bias in datasets where most features are assumed to be independent (*39*).

To search for plasma proteins displaying similar relationships with WLZ across both studies, the aptamer WLZ-associations from the MAM-phase (Eq. 15) and primary MAM study [**table S8** from (*7*)] were filtered to those that were present in the SomaScan assays used in both studies. The *β*_1_(Δ *protein*_*ij*_) *P*-values were inputted to CMA (*14, 15*) to generate consensus *P*-values and were subsequently controlled for FDR using a Benjamini-Hochberg correction (significance determined by q-value < 0.05). The direction of association was assigned by the sign of the *β*_1_(Δ *protein*_*ij*_) of Eq. 15 (positive or negative) and the sign of the published WLZ associations from the primary MAM study. Cases where the association coefficients were in opposite directions between the studies were common among poor fits and were not considered positively- or negatively-WLZ-associated (**table. S9A)**. We next applied an over-representation analysis of GO Biological Processes to the positively WLZ-associated protein group and the negatively WLZ-associated protein group with clusterProfiler (enrichGO, ont = ‘BP’, R package 4.6.2, q-value < 0.05) (*40*).

We analyzed the Eq. 12 and Eq. 13 *β*_*1*_(*treatment arm) P*-values using CMA to identify whether sets of WLZ-associated plasma proteins were enriched in the MDCF-2 or RUSF treatment groups across the MAM-phase of the current study and the previous RCT of children with primary MAM. The proteins were ranked by -log_10_(CMA *P*-value)*(direction of association). The direction of association indicated by the *β*_*1*_(*treatment arm*) coefficients of Eq. 12 and Eq. 13, and cases where the direction of association conflicted between the *β*_*1*_(*treatment arm*) coefficients for a particular protein were omitted. GSEA was performed using the “fgsea” function (R package fgsea v1.24.0, default parameters, q-value < 0.05) (*41*) on the list of proteins ranked by their treatment associations, and using the positively and negatively WLZ-associated plasma protein sets as enrichment groups. Leading edges proteins, defined within the fgsea analysis as those driving enrichment of a given set of features, were obtained from the GSEA results.

### Microbiome analyses

#### Isolation of nucleic acids from feces

Samples were prepared for shotgun sequencing in a manner similar to the previously reported trial (*9*) with further details provided in Supplemental Methods.

#### Analysis of 23 enteropathogens by multiplex qPCR

The qPCR assay was performed as previously described (*7*). Enteropathogen abundances were modeled for associations with WLZ using the following linear model:

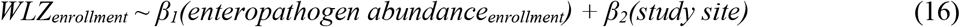

Kruskal-Wallis tests were applied to identify statistically significant (q-value < 0.05) differences in enteropathogen abundances between study sites and treatment arms at enrollment, baseline, and the end of MDCF-2 and RUSF treatment. A paired Wilcoxon signed-rank test was applied to test for differences in enteropathogen abundances before and after MDCF-2 or RUSF treatment.

#### MAG generation

MAG assembly and taxonomic assignments were performed in a similar manner to the previous study (*9*), and is described in detail in Supplemental Methods.

#### Principal components analysis (PCA) of MAG abundance profiles

PCA of MAG abundances was conducted using the ‘prcomp’ function in R. Unfiltered count data were first normalized using the implementation of the Variance Stabilizing Transformation (VST) in DESeq2 (*42*). VST-normalized data were analyzed using PCA after centering the data. Functions available in the ‘factoextra’ package v1.0.7 (*43*) were used to extract and analyze eigenvalues plus obtain data regarding the contributions of individual MAGs and projections of samples along each principal component explaining variance above a background threshold. To calculate this threshold, we randomly sampled the input matrix without replacement while retaining MAG and sample designations. Next, we performed PCA on the randomized matrix and set our background threshold as the variance explained by PC1 of this analysis.

#### MAGs associated with study site, treatment arm and sex at individual timepoints

We searched for VST-normalized MAG abundances associated with a study site, treatment group, or sex at enrollment, baseline, weeks 4, 8 and 12 of MDCF-2 and RUSF treatment, as well as the one-month follow-up timepoint using the following linear model for each MAG and timepoint:

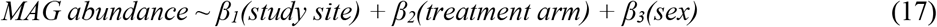

#### MAG diversity

Diversity metrics (both α- and β-diversity) were calculated in R using the vegan package v2.5-7 (*44*). Both the Shannon Index and Bray-Curtis Dissimilarity were calculated on TPM-normalized datasets filtered as described above. We modeled changes in alpha diversity and Bray-Curtis dissimilarity between enrollment and end of acute nutritional rehabilitation using linear mixed effects models:

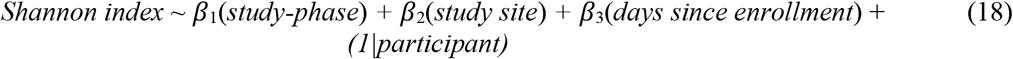

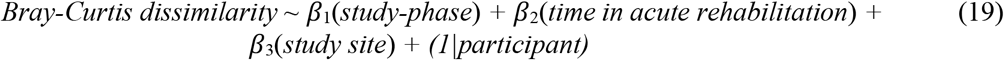

#### Identifying MAG abundances associated with time and study site during acute nutritional rehabilitation

We used the following linear mixed effects model using the dream function in the variancePartition R package, v1.24.0 (*45*) to identify the effects of acute nutritional rehabilitation on the abundances of individual MAGs represented in the fecal microbiomes of study participants:

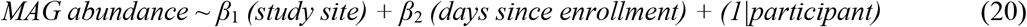

#### Identifying WLZ-associated MAGs

Linear mixed-effects models were used to relate the abundances of MAGs identified in each trial participant to WLZ. However, such analyses required different analytical strategies given the differences in fecal versus anthropometric sampling density in the SAM and MAM phases of the study. In the SAM acute rehabilitation phase, anthropometry data was available only at the beginning (enrollment) and end of this phase, whereas fecal samples were collected at 1-to-3-day intervals for each child, with the total number of fecal samples obtained determined by the length of time required for each child to achieve WLZ >-3. To analyze these datasets, we first calculated a rate of change for WLZ for each participant using the measurements taken at the beginning and end of the acute nutritional rehabilitation phase and the time spent in this phase. Next, we modeled the abundance trajectory of each of the 613 prevalent and abundant MAGs (defined as having >5 TPM in >40% of samples) in each participant by relating VST-transformed DESeq2 v1.40.2 (*42*) MAG abundances to the number of days after enrollment. Finally, we modeled the relationship between these two metrics using Eq. 1 (described in the main text). ANOVA was used to determine the significance (*P-*values) of the relationship between the MAG abundance trajectories and rate of change of WLZ and corrected to control False Discovery Rate using the Benjamini-Hochberg method. MAGs with statistically significant WLZ associations in the SAM phase of the study were identified as those having q-values < 0.05.

We filtered the VST-transformed abundance dataset to include only WLZ-associated MAGs identified in the SAM (Eq. 1) or MAM phase (Eq. 2), and then employed the following linear model to examine the relationship between the rate of anthropometric recovery from SAM and the abundances of MAGs at the time of enrollment:

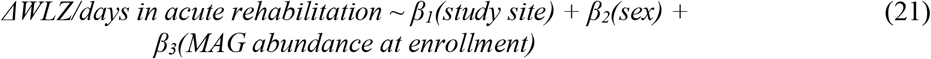

MAGs displaying a *β*_*3*_ coefficient with an FDR-corrected q-value < 0.05 were deemed ‘significantly predictive’.

In the MAM phase of the study, we employed linear mixed effects models for each MAG to relate VST-transformed MAG abundances to WLZ over time while controlling for study site and treatment group (see Eq. 2 in the main text). ANOVA was used to determine the significance of the interaction of MAG abundance and study week with WLZ and then corrected for multiple testing (q < 0.1). We also searched for significant associations (q-value <0.1) between the abundances of MAGs that were positively correlated with WLZ using the following linear mixed effects model, where *i* and *j* represent MAG IDs (these IDs are described in **table S13D**).

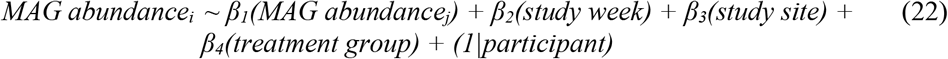

#### Impact of MDCF-2 and RUSF treatment on MAG abundance across timepoints

Linear mixed effects models were implemented to identify treatment-specific changes in VST transformed MAG abundances during the MAM phase while controlling for differences in MAG abundance between the Dhaka and Kurigram study sites.

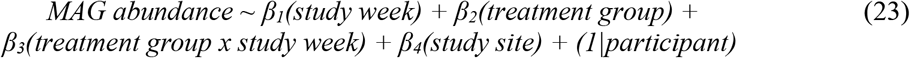

We used ANOVA to determine the significance of the interaction of treatment group and study week with MAG abundance and controlled for multiple testing using FDR. Significant relationships were designated with a q-value < 0.1.

#### Predictions of anthropometric responses based on MAG abundances at enrollment and at baseline

We employed the model described in Eq 23. to determine whether projection of the microbial community configuration of a given fecal sample along PC1, PC2 or PC3 at enrollment or baseline was predictive of ponderal growth response using the following model:

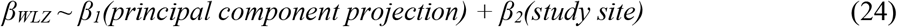

A linear model was applied to relate the abundance of each *MAG*_i_ at enrollment or baseline to the rate of change of WLZ for each participant while controlling for study site. Significance was determined by ANOVA (q-value <0.1).

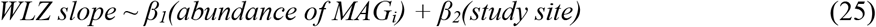

#### Annotation of antibiotic resistance markers

We used AMRFinderPlus v3.11.2 (*46*) together with the corresponding AMRFinderPlus database (version 2022-12-19.1) to identify antimicrobial resistance genes in MAG assemblies. We employed these annotations, the log_2_ transformed fold-changes of each MAG abundance calculated using Eq. 20, and GSEA (fgsea v1.24.0) (*41*) to identify enriched classes of antimicrobial resistance genes in MAGs whose abundances increased during acute nutritional rehabilitation.

#### Identification of metabolic phenotypes in WLZ-associated MAGs

We first filtered the 106 metabolic pathways to those present in more than 5% of the 613 prevalent and abundant MAGs. Next, we related the presence or absence of the resulting 90 metabolic phenotypes to the coefficient of each MAG association with WLZ [defined from Eq. 2 as *β*_*3*_(*study week:MAG abundance*)] as described in Eq. 3.

#### Comparison of carbohydrate utilization pathways between WLZ-associated and WLZ-non-associated P. copri MAGs

A contingency table was created using the 18 carbohydrate utilization pathways that were significantly associated with WLZ and present in the 33 *P. copri* MAGs. The first row of the table consisted of the group of WLZ-associated *P. copri* MAGs. The second row contained the group of non-WLZ-associated *P. copri* MAGs. The columns represented the 18 different pathways. Thus, each cell in the table represented the number of pathway occurrences in each group of *P. copri* MAGs. A Fisher’s exact test was employed to determine if the composition of pathways was significantly different between groups (*P*-value < 0.05).

#### Comparison of WLZ-associated carbohydrate utilization pathways between P. copri and other Prevotella MAGs

This analysis used the 19 carbohydrate utilization pathways associated with WLZ in the 63 ‘prevalent and abundant’ *Prevotella* MAGs. A contingency table was created where the first row contained *P. copri* MAGs, the second row contained all *Prevotella* MAGs not assigned to *P. copri*, and the columns represented the 19 carbohydrate utilization pathways. A Chi-squared test was used to determine if the composition of pathways was significantly different between the two groups of MAGs (*P*-value < 0.05).

#### Statistical Information

Statistical analyses were conducted using the approaches described in *Methods* and in the figure legends. Sample sizes are indicated along with each statistical test. All statistical unless otherwise specified. All measurements were performed using distinct samples. Statistical significance reported as *P-*values is uncorrected, while significance reported as q-values indicates correction to control false discovery rate using the Benjamini-Hochberg approach. For anthropometric measures, analyses are reported for the flood-unaffected trial participants who completed the trial per protocol. The results of analyses encompassing flooding unaffected participants, flooding unaffected and affected, and the intent-to-treat analyses are tests are two-tailed reported in **table S6**.

## Supporting information

Suplemental tabes

## Data Availability

All data produced in the present study are available upon reasonable request to the authors and metagenomic sequencing data will be available in the European Nucleotide Archive under accession PRJEB63587.

https://gitlab.com/sjhartman/post-sam-mam-code-repository

https://github.com/rodionovdima/PhenotypePredictor

## SUPPLEMENTARY MATERIALS

**Supplementary Methods**

**Figs. S1 to S6**

**Tables S1 to S17**

**References (*47-71*)**

## Acknowledgements

We thank the Bangladeshi study participants and their families for their participation, plus the dedicated staff of the icddr,b for conducting the study, Martin Meier, Justin Serugo, Su Deng, Kazi Ahsan, Jessica Hoisington-Lopez, and MariaLynn Crosby for superb technical assistance, members of the Genome Technology Access Center at Washington University School of Medicine for Illumina NovaSeq-based generation of fecal DNA shotgun sequencing datasets, and Eric Martin and Brian Koebbe for high-performance computing support.

## Author Contributions

I.M., under supervision of T.A. and with the assistance of N.N.N., Md.M.I., S.Huq, M.Z., M.M., T.I., K.M. and icddr,b and Terre des Hommes workers conducted the clinical study, including collection of all of the metadata and biospecimens used for the analyses described in this report. M.J.B. oversaw databases of clinical metadata and the biospecimen archive and contributed to various aspects of data analysis. M.C.H., I.M., and R.Y.C. analyzed anthropometry data. S.J.H. analyzed the plasma proteome with guidance from V.M. and M.A.P. D.M.W. helped prepare short-read shotgun sequencing datasets and aided in linear modeling strategies. S.J.H. and M.C.H assembled MAGs, identified those that were WLZ-associated and conducted analyses of the enrichment of their encoded functions. D.A.R. and A.L.O performed metabolic pathway annotations of MAGs while S. Henrissat, B.H., and N.T. annotated CAZymes and PULs in MAGs. T.A. and J.I.G. oversaw this research. S.J.H., M.C.H., M.J.B., and J.I.G. wrote the paper with invaluable input from co-authors.

## Data and Materials Availability

Shotgun DNA sequencing datasets generated from fecal samples plus annotated MAG sequences are available in the European Nucleotide Archive under accession PRJEB63587. Code detailing the steps in the MAG assembly workflow, analysis and plasma protein analysis datasets are available from gitlab (https://gitlab.com/sjhartman/post-sam-mam-code-repository). Code for annotation of bacterial genes and prediction of metabolic pathway presence/absence is available from GitHub (https://github.com/rodionovdima/PhenotypePredictor). Human biospecimens collected from Bangladeshi children are the property of icddr,b. Material Transfer Agreements between icddr,b and Washington University are in place for use of these samples. Requests for materials should be made to J.I.G.

