## Supplementary material for "A microbiome-directed therapeutic food for children recovering from severe acute malnutrition": Suplemental Material and Methods

**Supplementary Methods**

***Quantification of plasma proteins*** - Plasma samples were collected using the same methods described in our previous RCT of MDCF-2 treatment in children with primary MAM (*7*) and at the following time points: enrollment, baseline, week 4 and week 12 of the MAM treatment phase, and at the conclusion of the 1-month post-treatment follow-up. Samples were analyzed using SomaScan 7K Proteomic Assay plasma/serum kit (*47*). A total of 267 of the 7,288 Slow Off-rate Modified Aptamer reagents (aptamers) targeting human proteins were flagged and removed from subsequent analyses due to their failure to meet quality control thresholds. Forty-three aptamers with a mean abundance within three standard deviations of the mean value obtained with a blank (buffer alone) control sample were also removed, yielding 6,978 aptamers representing 6,138 unique proteins. Plasma protein abundances were log transformed and quantile normalized for all downstream analyses.

***Isolation of nucleic acids from feces*** **-** Fecal samples were homogenized with mortar and pestle under liquid nitrogen approximately 50 mg of each homogenized sample was transferred to a 2 mL screw top tube (Axygen, SCT-200-SS-C-S) along with one 3.97 mm steel ball and 250 µL of 0.1 mm zirconia/silica beads and (iii) a 500 µL mixture of 25:24:1 parts phenol:chloroform:isoamyl alcohol (pH 7.8-8.2), 210 µL of 20% SDS, and 500 uL of 2X buffer A (200 mM NaCl, 200 mM Trizma base, 20 mM EDTA) was added to each tube. Samples were then subjected to bead-beating for 4 minutes in a Biospec Minibeadbeater-96, followed by centrifugation at 3220 x g for 4 minutes. A 100 µL fraction of the resulting aqueous phase was transferred to a deep 96-well plate together with 70 µL isopropanol and 10 µL 3M NaOAc, pH 5.5; the solution was subsequently mixed by pipetting 10-times. The crude DNA mixture was chilled at -20 °C for approximately 1 hour to precipitate the nucleic acids and then centrifuged at 3,220 x g at 4 °C for 15 minutes before removing the supernatant, yielding nucleotide-rich pellets. A Biomek FX robot was used to add 300 µL Qiagen Buffer RLT to the pellets and to resuspend the DNA by pipetting up and down 50-times. A 400 µL aliqout of the resulting mixture was transferred to an AllPrep 96 DNA plate (Qiagen, Catalog # 80311) which was centrifuged at 3,220 x g for 1 minute at room temperature. DNA was then eluted from the column and retained.

DNA extracted from 760 fecal samples was used to prepare shotgun sequencing libraries [reduced-volume protocol based on the Nextera XT (Illumina) strategy (*48*)]. Libraries were quantified, balanced, pooled and sequenced (Illumina NovaSeq 6000, S4 flow cell) to a depth of $2.2 \times 10^7 \pm 2.9 \times 10^6$ 150 nt paired-end reads/sample (mean ± SD). DNA reads were demultiplexed (bcl2fastq, Illumina), trimmed to remove low quality bases, and processed to remove read-through adapter sequences Trim Galore v0.6.4 (*49*). Read pairs where the length of either read was <50 nt after quality and adapter trimming were discarded. The remaining reads were mapped to the human genome (UCSC hg19) using bowtie2 v2.3.4.1 (*50*) and were filtered to remove *H. sapiens* sequences.

***MAG assembly*** - Preprocessed, short-read shotgun data were aggregated within each flooding-unaffected participant's fecal sample set (n=9-14 samples/participant; 70 participants) prior to MAG assembly. This strategy was adopted to enable the contig abundance calculations required by the MAG assembly algorithms employed below, while limiting the pitfalls of aggregate assembly across individuals. Metagenomic assemblies were generated from each dataset obtained from 70 participants using MegaHit v1.1.4 (*51*). The resulting contigs were quantified in each assembly by mapping reads from each of the corresponding participant's fecal samples to the assembled contigs with kallisto v0.43.0 (*52*). Contigs were assembled into MAGs using MaxBin2 v2.2.7 (*53*) and MetaBAT2 v2.12.1 (*54*). The results of both MAG assembly strategies were merged and dereplicated using DAS Tool v1.1.2 (*55*) on a per-participant basis.

MAGs were assessed for completeness and contamination ['lineage_wf' command in CheckM, v1.1.3; (*56*)] and refined ('tetra', 'outliers', and 'modify' commands) to remove contaminating contigs. Additional refinement based on the distribution of phylogenetic markers present in each MAG was performed ['phylo-markers', 'clade-markers', and 'clean-bin' commands in MAGpurify v2.1.2 (*57*) resulting in 3,592 MAGs, or 51.3±12.9 (mean±SD) MAGs/participant. We subsequently employed a stringent (≥ 90% complete, ≤ 5% contaminated, ANI ≥ 99%) bulk dereplication (options '-l 50000', '–completeness 90', '–contamination 5', '-pa 0.9', '-sa 0.99') in dRep v2.3.2 (*58*) that yielded 754 nonredundant MAGs across all participants' fecal samples. MAG assembly summary statistics were collected from CheckM and quast v4.6.1 (*59*) analyses, and aggregated (**table S11A**). MAGs were initially annotated using prokka v1.14.0 (*60*). To quantify MAG abundances, a single kallisto quantification index was created from the dereplicated MAGs. Reads from each fecal DNA sample were then mapped to this index to quantify the abundance of each of the 754 MAGs in each sample (*61*).

***Assigning taxonomy to MAGs* –** Taxonomic assignments were initially made using the Genome Taxonomy Database Toolkit v1.5.0 (GTDB-Tk) (*27*) and corresponding database (release 202). For phylogenetic analysis of *P. copri* isolates and MAGs, or *Gemmiger* MAGs, we first used CheckM v1.1.3 (*56*) to extract and align the amino acid sequences of 43 single copy marker genes in each isolate or each MAG, plus reference isolate genomes of each genus as indicated in **Fig. 4A.** Published reference genomes of (i) cultured members of the genus *Prevotella,* including the genomes previously reported (*28*) for clade typing, as well as (ii) *G. formicilis*, *G. qucibialis*, and *G. gallinarum*. For each genome set, concatenated marker gene sequences were analyzed using fasttree v2.1.10 (*62*) to construct a phylogenetic tree using the Jones-Taylor-Thornton model and CAT evolution rate approximation, followed by tree rescaling using the Gamma20 optimization. The tree was processed using ape v5.6-2 (*63*) to root each tree with an appropriate reference genome as an outgroup. Trees were plotted using ggtree v3.2.1(*64*). Genome similarities between reference isolates and MAGs were quantified by calculating the average nucleotide similarity (ANI) score with dRep ['compare' method, v0.2.10 (*58*)].

***Annotation of metabolic pathways represented in MAGs* -** MAG metabolic pathway annotations were performed as described in a previous publication (*9*). Briefly, our strategy employed a combination of public domain tools, custom code and a reference collection of functionally annotated genomes from the mcSEED database encompassing *in silico* reconstructions of major metabolic pathways in 2,856 reference bacterial genomes representing members the human gut microbiota (*30*). The mcSEED database is based on the subsystem concept (*29*), in which each metabolic subsystem is composed of a set of gene functions (enzymes, transporters, and transcriptional regulators) that contribute to metabolic pathway variants (*65*) involved in utilization and/or catabolism of major nutrients (carbohydrates, oligosaccharides, and amino acids), biosynthesis of vitamins/cofactors and amino acids and generation of fermentation end-products (e.g., short-chain fatty acids) (*66–68*).

Protein-coding genes in the 754 input MAGs were identified using Prokka (v1.14, --metagenome flag) (*60*). These proteins were aligned [Diamond v2.0.15, default parameters, (*69*)] with proteins in genomes comprising the mcSEED reference database. This effort yielded 224,867 functionally annotated proteins across the 754 MAGs (**table S15B**); these proteins represented 1,482 nonredundant functional roles from 80 curated mcSEED subsystems. The annotated MAGs and alignments to 2,856 reference MAGs were inputted to the three following complementary approaches to predict MAG metabolic 'phenotypes': (i) direct application of binary phenotypes (presence or absence of complete metabolic pathways) from reference genomes determined from phylogenetic Neighbor Groups (NG); (ii) interpretation of patterns of gene presence/absence using explicit Pathway Rules (PR) and (iii) interpretation of gene presence/absence patterns using pathway-specific Random Forest machine learning (ML) models trained on the contents of the mcSEED reference database (see (*9*) for details of these approaches). The binary phenotype assignments across all three methods were used to produce a consensus binary phenotype table for each of 106 functional metabolic pathways over the entire set of 754 MAGs (**table S15C**). Figures depicting the presence and absence of metabolic phenotypes in a circularized layout were generated with the circos.heatmap function from the R package circlize v0.4.15 (*70*).

***Annotation of Carbohydrate-active enzymes (CAZymes) and Polysaccharide Utilization Loci (PULs)* –** CAZymes were annotated following the CAZy classification scheme (*71*). To do so, amino acid sequences from MAGs were analyzed using a bioinformatic workflow that performs homology searches between each MAG's protein coding sequences and the CAZy database (*72*). This approach specifically accommodates the modular structure of CAZymes, which often carry a variable number of both ancillary modules and catalytic domains. Details of this workflow and its application are provided in a previous publication (*73*). To predict the specificity of each CAZyme in *G. formicilis* and *P. copri* genomes, a similarity search of the protein sequences was performed using BlastP against a library of CAZymes with experimentally determined activities (*72*). Each annotated CAZyme in *G. formicilis* and *P. copri* was assigned the activity of the best homolog match from the experimentally characterized library.

PULs were identified in *Prevotella* MAGs by combining information from marker genes (SusC/SusD pairs), operon structure and CAZyme annotations (*32*). The experimentally validated substrate specificities of homologs of CAZymes contained in each PUL were used to infer its glycan target(s). We compared the conservation of PULs between *P. copri* MAGs identified in the current study to *P. copri* MAGs and cultured Bangladeshi isolates identified in our prior study of MDCF-2 in children with primary MAM, using an approach described previously (*9*). Briefly, we compared the presence/absence and sequence similarity of genes in each PUL in each MAG with the corresponding homologous PULs in Bg0019 MAG (the *P. copri* MAG that had exhibited the strongest association with WLZ in Bangladeshi children with primary MAM). We used this information to classify eachasd f PUL as 'conserved', 'structurally distinct' or 'not conserved/absent' compared to MAG Bg0019. These patterns of PUL conservation for each MAG were used, together with predictions regarding the substrate specificities of each PUL, as additional criteria to interpret the predicted functional similarity of *P. copri* MAGs from the current study to *P. copri* MAGs and isolates identified the previous study.

### Supplementary Figures:

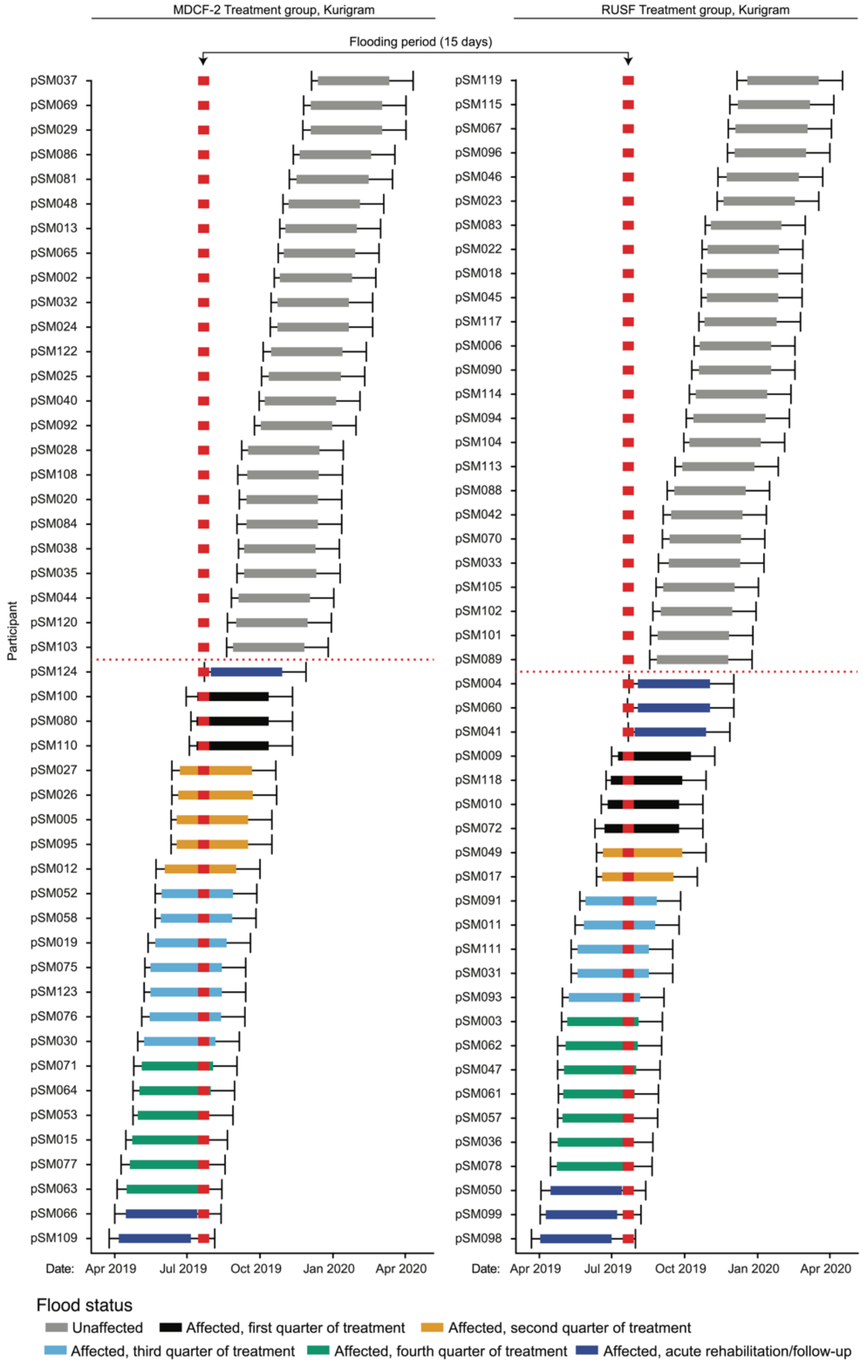


**fig. S1 – Timing of flooding in Kurigram relative to the stage of participation in the trial.** Each thick horizontal bar indicates the period of MDCF-2 or RUSF treatment during the MAM phase for participants in the Kurigram study site. The thin black bar for each participant extends to the left to indicate the time of enrollment through the end of the acute rehabilitation phase, and to the right to indicate the end of the one-month post-intervention follow-up period. The 15-day period of flooding is denoted by the red bar. Trial participants were classified based on whether the flood occurred during (i) the acute rehabilitation phase for SAM, (ii) the 90-day treatment period with MDCF-2 or RUSF, (iii) the post-intervention period, or (iv) not at all during any period of the trial. These latter ‘flood unaffected’ Kurigram children were included in our analysis (n=49) and are indicated by the thick gray bars

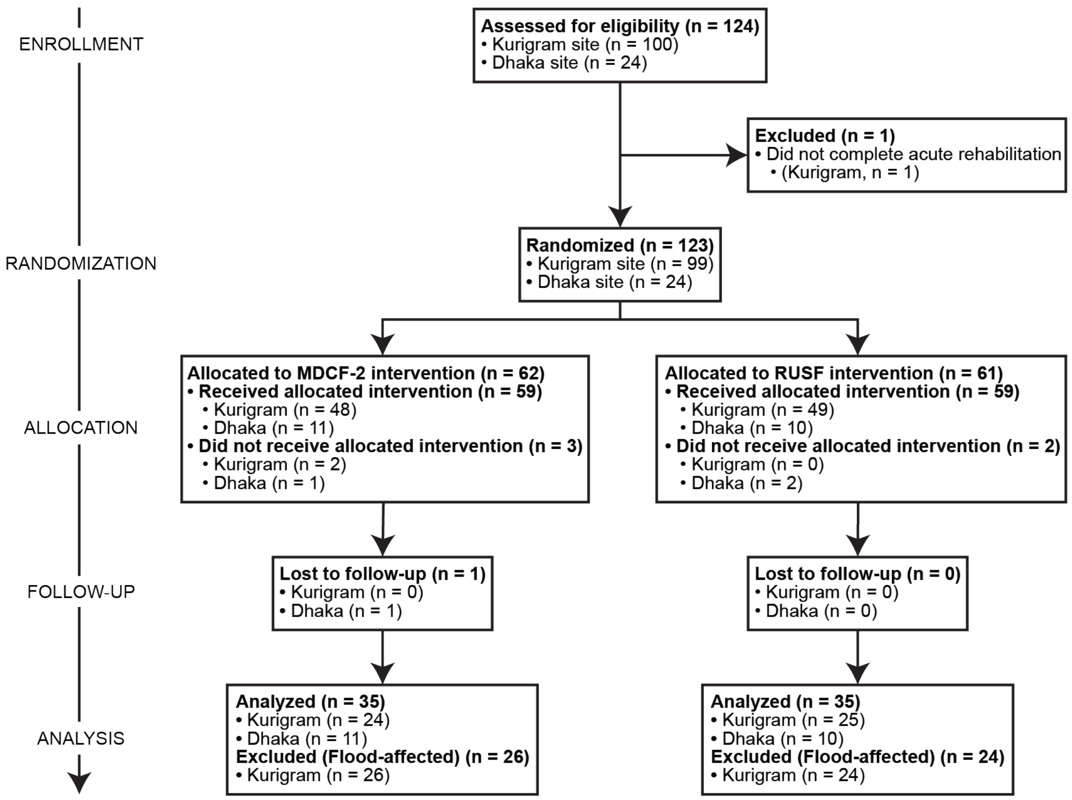


**fig. S2 –Consort diagram**. Schematic representing the number of participants considered at each step of the trial according to treatment group, study site, adherence to the trial protocol and effect of flooding.

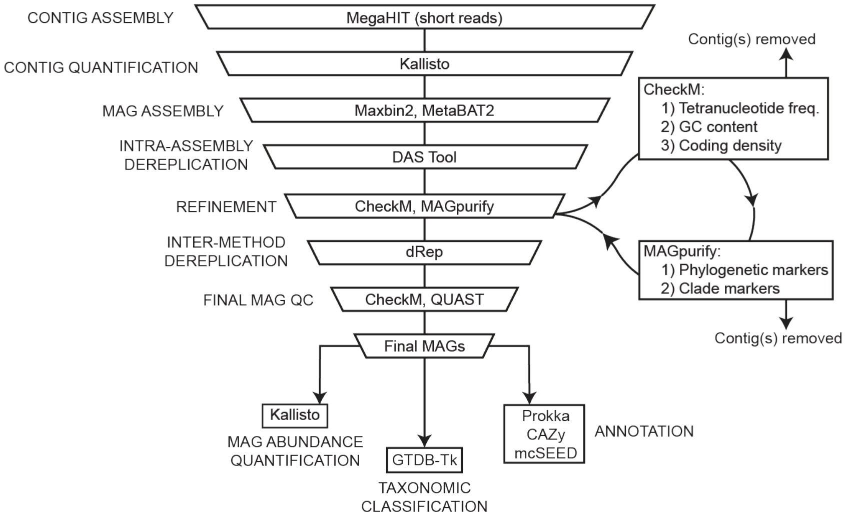


**fig. S3 –Schematic of procedures used for MAG assembly, quality control, functional annotation, taxonomic classification, and quantification of abundance**.

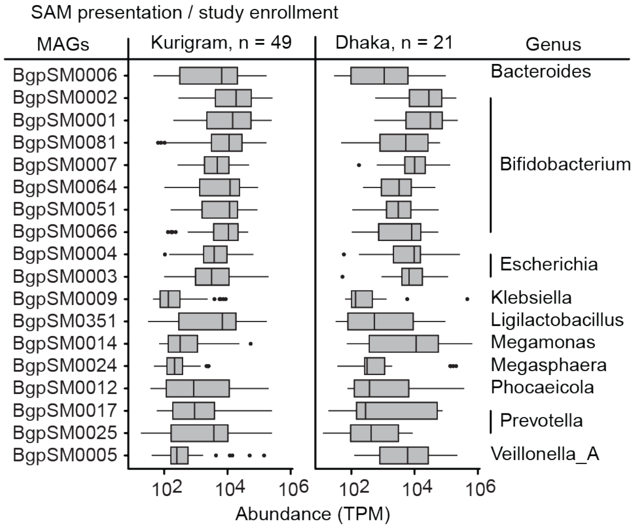


**fig. S4 – Abundances of the 10 most abundant MAGs in the fecal microbiomes of children with SAM sampled at the time of enrollment at each site.** Boxplots show the median, first and third quartiles; whiskers extend to the largest value within 1.5 × the interquartile range.

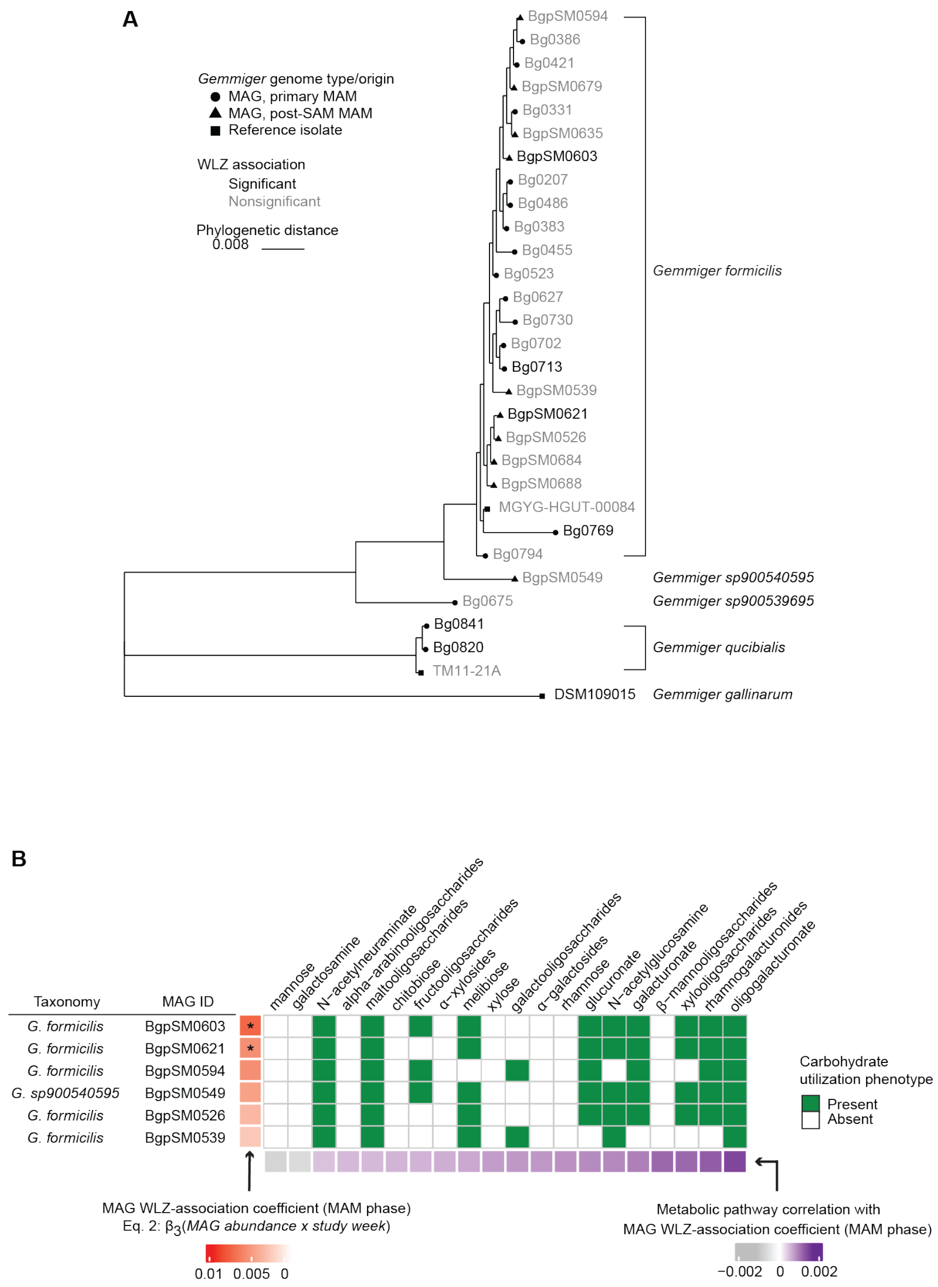


**fig. S5 – Analysis of phylogenetic relationships and the representation of carbohydrate utilization pathways in *Gemmiger* MAGs**. (**A**) Marker gene-based phylogenetic tree indicating the inferred relationships between *G. formicilis* and related *G. qucibialis* MAGs from the current study, our prior study of children with primary MAM and reference *Gemmiger* isolate genomes. WLZ-associated MAGs from either study are colored black. The GTDB taxonomic assignment of each MAG or isolate is depicted on the right. **(B)** Carbohydrate utilization pathways present in two significantly positive WLZ-associated *G. formcilis* MAGs from the current study compared to the three other *G. formicilis* and *G.* sp900540595 MAGs prevalent and abundant but not WLZ-associated. MAGs are ordered from top to bottom by the strength of their WLZ association during the MAM phase. All of the metabolic pathway correlations with WLZ-association coefficients shown in the bottom row are statistically significant.

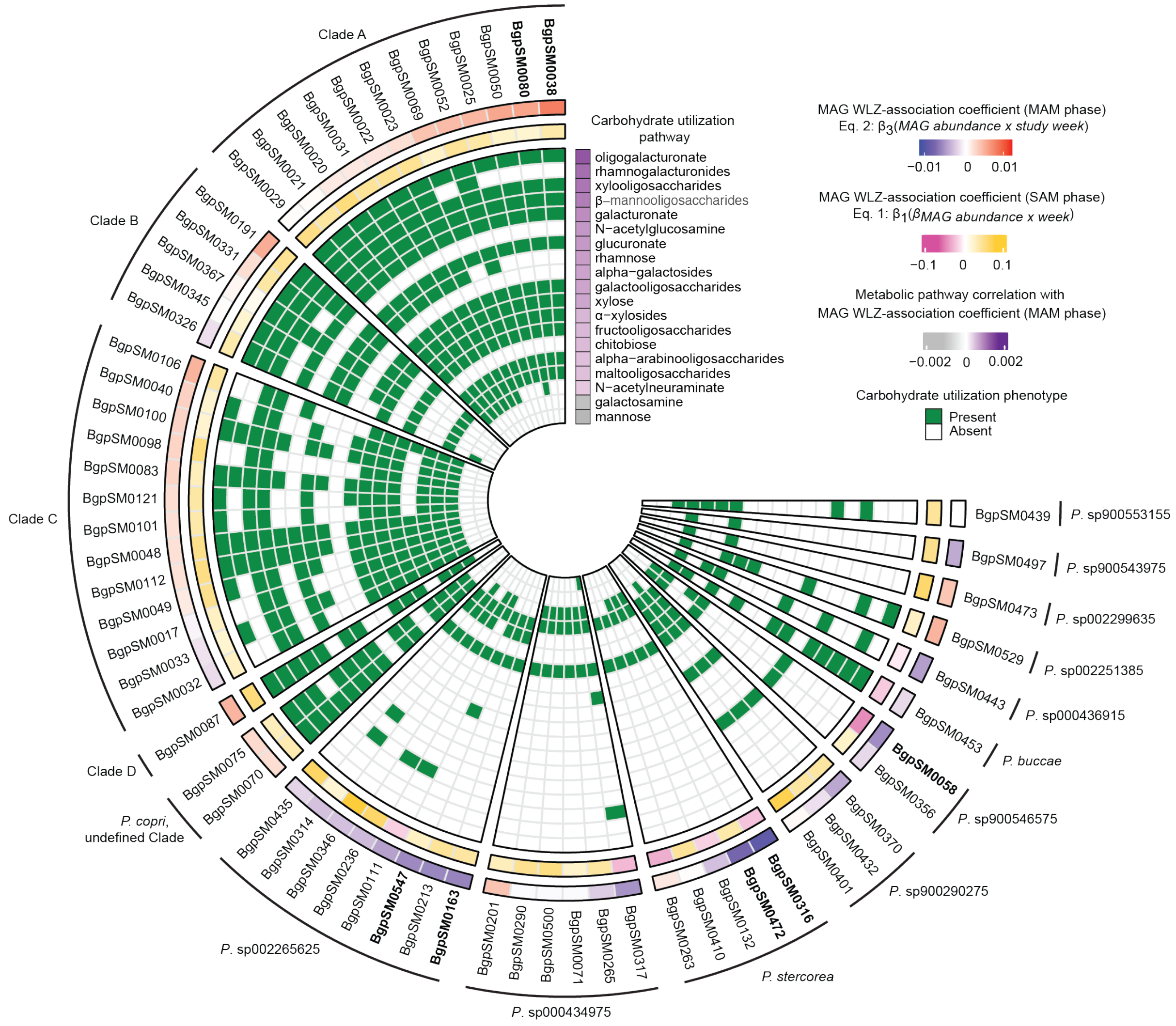


**fig. S6 – Representation of the presence or absence of carbohydrate utilization pathways in *Prevotella* MAGs from the current study.** The MAM phase WLZ association of each MAG is shown in the outermost row and the SAM phase WLZ association in the second to outermost row. MAGs are grouped by species or by clades if they were assigned to *P. copri.* Bolded MAGs are significantly associated with WLZ in the MAM-phase.

**Supplementary Tables:**

**Table 1. Enrollment and baseline characteristics and samples obtained from children in the study.** (**A**) Anthropometry data. (**B**) Ledger of fecal and plasma samples. (**C**) Demographic and environmental conditions at enrollment.

**Table 2. Effect of flooding on the anthropometry of study participants residing in Kurigram during MDCF-2 or RUSF treatment.**

**Table 3. Enrollment and baseline characteristics of participants at each study site.** (**A**) Enrollment and baseline anthropometric characteristics for all participants. (**B**) Enrollment and baseline anthropometric characteristics for flood-unaffected participants in Kurigram. (**C**) Enrollment and baseline characteristics separated by treatment group within each study site for all participants. (**D**) Enrollment and baseline characteristics separated by treatment group within each study site for flood-unaffected participants in Kurigram.

**Table 4. Anthropometric comparisons at baseline between the post-SAM MAM and primary-MAM studies. (A)** Difference in WLZ between -SAM MAM and primary-MAM studies at baseline. **(B)** Difference in WAZ between post-SAM MAM and primary-MAM studies at baseline. **(C)** Difference in LAZ between post-SAM MAM and primary-MAM studies at baseline. **(D)** Difference in MUAC between post-SAM MAM and primary-MAM studies at baseline.

**Table 5. Food frequency questionnaire responses and therapeutic food consumption during the 3-month period of treatment for MAM.** (**A**) Percent of MDCF-2 and RUSF consumed throughout the trial. (**B**) Associations of food frequency questionnaire responses with treatment arm across time during the MAM phase. (**C**) Associations of food frequency questionnaire responses with treatment group and study site at enrollment. (**D**) Associations of food frequency questionnaire responses with treatment group and study site at baseline. (**E**) Associations of food frequency questionnaire responses with treatment group and study site at the end the MAM-phase.

**Table 6. Clinical response to MDCF-2 compared to RUSF supplementation.** (**A**) Intent-to-treat analysis: Anthropometric measures of growth during nutritional supplementation of all participants, including those who did not complete the trial. (**B**) Anthropometric measures of growth during nutritional supplementation of flood-affected and flood unaffected participants. (**C**) Anthropometric measures of growth during nutritional supplementation of flood-unaffected participants. (**D**) Intent-to-treat analysis: changes in anthropometric measures of growth during MDCF-2 or RUSF supplementation and the one-month follow-up period for all participants, including those who did not complete the trial. (**E**) Anthropometric measures of growth during supplementation and the one-month follow-up period for flood affected and unaffected participants. (**F**) Anthropometric measures of growth during nutritional supplementation and the one-month follow-up period for flood unaffected participants.

**Table 7. Comorbidity analysis.** (**A**) Association between the occurrence of comorbidities and the change in WLZ during the MAM phase in flood-unaffected children. (**B**) Associations of comorbidity frequencies between MDCF-2 and RUSF over time during the MAM phase in flood-unaffected children.

**Table 8. Plasma protein associations with WLZ.** (**A**) Associations between levels of plasma protein, treatment group and study site in the MAM-phase. (**B**) Relationship of plasma protein levels with WLZ during the SAM and MAM treatment phases.

**Table 9. Correlated meta-analysis of plasma protein associations with WLZ and treatment groups** (**A**) Correlated meta-analysis of plasma protein WLZ associations in the MAM phase of the current study and in the previous primary MAM study. (**B**) Over-representation analysis of GO Biological Processes among positively WLZ-associated proteins from the correlated meta-analysis. (**C**) Over-representation analysis of GO Biological Processes in negatively WLZ-associated proteins from the correlated meta-analysis. (**D**) Correlated meta-analysis of MAM phase of the current study and the previous primary MAM study plasma protein associations with treatment. (**E**) Enrichment of WLZ-associated plasma proteins in the MDCF-2 or RUSF treatment groups. (**F**) Overlap of proteins significantly associated with WLZ during the SAM phase and WLZ during MAM, as identified by the correlated meta-analysis.

**Table 10. qPCR-based analysis of enteropathogen abundances in fecal samples.** (**A**) Analysis of enteropathogen abundance between study sites at enrollment. (**B**) Analysis of enteropathogen abundance between study sites and treatment groups at baseline. (**C**) Analysis of enteropathogen abundance between study sites and treatment groups at the end of the MAM phase. (**D**) Analysis of the change in enteropathogen abundance from the beginning to end of the MAM-phase between study sites.

**Table 11: Summary of MAG assembly statistics, abundances, and diversity.** (**A**) MAG assembly statistics and taxonomic assignments. (**B)** MAG abundances. (**C**) MAG-based microbiome diversity metrics.

**Table 12: Principal components analysis of MAG abundances over the course of the SAM and MAM treatment phases of the trial.** (**A**) Variance explained by each principal component. (**B**) MAG drivers of variation among the three principal components explaining the most variance.

**Table 13: Analysis of MAG abundances over time and the relationship between MAG abundances and presence of antibiotic resistance markers.** (**A**) Changes in MAG abundance during acute rehabilitation for SAM. (**B**) MAG abundance associations with study site, treatment arm, and sex at each fecal sampling timepoint. (**C**) Annotation of antibiotic resistance markers according to AMRFinderPlus. (**D**) Enrichment of predicted antibiotic resistance traits in MAGs according to their abundance response during acute nutritional rehabilitation for SAM.

**Table 14: Associations between changes in the abundances of MAGs and rates of ponderal growth.** (**A**) Association between the abundance of each MAG and WLZ in the SAM treatment phase. (**B**) GSEA of taxa in MAGs ranked by their association with WLZ in the MAM treatment phase. (**C**) Association of the abundance of each MAG with WLZ in the MAM treatment phase. (**D**) Association between the abundances of positively WLZ associated MAGs during the MAM treatment phase. (**E**) Changes in MAG abundance during MDCF-2 versus RUSF treatment for MAM.

**Table 15: Predicted metabolic pathways in each MAG and the relationship between pathway representation and the WLZ association of each.** (**A**) Metabolic pathway (phenotype) descriptions. (**B**) Gene annotations in metabolic pathways. (**C**) Metabolic pathway presence or absence in 754 high-quality MAGs. (**D**) Association of pathway presence with MAG-WLZ association.

**Table 16: PUL annotations of WLZ-associated *P. copri* MAGs.** (**A**) Analysis of PUL conservation compared to the WLZ-associated *P. copri* MAG Bg0019 from the primary MAM trial. (**B**) PUL annotations.

**Table 17: CAZymes present in positively WLZ-associated MAGs with predicted activity against xylooligosaccharides, maltooligosaccharides, glucuronate, galacturonate and oligogalacturonate.**
